# Cortical oscillations predict auditory grouping in listeners with and without hearing loss

**DOI:** 10.1101/2025.09.02.25334927

**Authors:** Nour Alsabbagh, Bob McMurray, Timothy D Griffiths, Joel I Berger, Kyogu Lee, Phillip E Gander, Inyong Choi

**Affiliations:** Dept of Communication Sciences & Disorders, The University of Iowa, Iowa City, USA; Dept of Otolaryngology, University of Iowa Hospital and Clinics, IA, USA; Dept of Psychological & Brain Sciences, University of Iowa, IA, USA; Dept of Neurosurgery, University of Iowa Hospitals and Clinics, IA, USA; Bioscience Institute, Newcastle University, UK; Dept of Intelligence and Information, Seoul National University, Seoul, South Korea; Dept of Radiology, University of Iowa Hospitals and Clinics, IA, USA

**Keywords:** Auditory grouping, stochastic figure-ground, sensorineural hearing loss, cochlear implant, hearing aid, cortical evoked responses, cortical oscillations

## Abstract

Auditory grouping relies on the ability to bind tones with coherent spectral features over time to form auditory objects. Sensorineural hearing loss (SNHL) degrades spectral resolution, and the extent of this degradation varies with the listening configuration. However, it remains unclear how SNHL impacts auditory grouping and whether different listening configurations affect this ability. This study investigated task performance and cortical activity during auditory object detection in four groups with different listening configurations: Twenty normal-hearing (NH) listeners, seventeen bilateral hearing aid users with acoustic-only stimulation (A-only), thirty-one cochlear implant (CI) users with acoustic and electric stimulation (A+E), and seventeen bilateral CI users with electric-only stimulation (E-only). While electroencephalography was recorded, participants performed a stochastic figure-ground task requiring the detection of spectrally and temporally coherent tone pips embedded in a background of random-frequency tone clouds. All groups achieved above 80% accuracy, though CI groups showed poorer performance compared to NH and A-only groups. Compared to NH listeners, the object-related evoked responses were weaker in A-only listeners and absent in CI groups. Delta (2-3.5Hz) and theta (4-7Hz) event-related synchronization (ERS) to the auditory objects were only observed in the NH group, except for the A+E group, which showed a delta ERS. However, all groups exhibited alpha (8-15Hz) and beta (17-30Hz) event-related desynchronization (ERD), with no significant group differences. Notably, individual differences in alpha and beta ERD predicted task accuracy. These findings suggest that alpha and beta cortical activity, measured during an auditory object detection task, reflects auditory grouping in any listening configuration.

## Introduction

Communication in daily life often involves listening to speech in challenging conditions such as in background noise, in a reverberant room, or when speech is quiet. Speech in noise is perhaps the most salient of these conditions, and one that presents a significant struggle for older adults and people with hearing loss.

The ability to segregate speech from noise relies on detecting auditory components with similar acoustic features and binding them across time and frequency to form auditory objects that can then be separated from the background (Griffiths & Warren, 2004). Auditory object formation involves a hierarchy that starts with peripheral encoding of acoustic signals at the level of the cochlea and ends with cognitive processes that occur outside the auditory cortex (Griffiths & Warren, 2004). This process is pre-attentive (O’Sullivan et al., 2015; Winkler et al., 2005); however, the formation of auditory objects can be enhanced with directed attention (O’Sullivan et al., 2015; Shamma et al., 2011).

A critical bottleneck in the hierarchy of auditory object formation is the fidelity of the cochlea in encoding the spectral and temporal features of the sound. This multidimensional representation of the auditory object then propagates along the auditory pathway to the auditory cortex (Shamma et al., 2011). However, if this representation lacks fidelity, it can hamper the ability to discover common auditory features that are the basis of object binding, which is essential for speech-in-noise (SiN) perception (Holmes & Griffiths, 2019; Holmes et al., 2021).

It is well known that people with sensorineural hearing loss (SNHL) experience impaired cochlear spectral resolution (Larsby & Arlinger, 1999; Turner et al., 1999). This is especially true for people with cochlear implants (CI) (Friesen et al., 2001; Henry & Turner, 2003), who rely on a small number of electrodes to code frequency information. This degradation in spectral resolution impairs SiN perception in people with SNHL (Arlinger & Dryselius, 1990; Ter Keurs et al., 1993) and CI devices (Friesen et al., 2001; Oxenham & Kreft, 2014). Simulation studies in normal hearing (NH) listeners that presented sounds that simulated CI processing, thereby reducing spectral resolution, have documented similar SiN impairments (Baer & Moore, 1993, 1994; Qin & Oxenham, 2003; Xu et al., 2020). Less well understood is how intermediate processes, such as auditory grouping, are disrupted by poor spectral resolution and may contribute to SiN deficits.

Here, we examined auditory grouping abilities in subjects with different listening configurations using the stochastic figure-ground (SFG) paradigm. This paradigm was developed to evaluate auditory grouping abilities that depend on temporal coherence across frequency bands (Teki et al., 2011, 2013, 2016). In this paradigm, the stimuli are composed of several tonal elements distributed across time and frequency (Figure 1). When auditory components are randomized across frequency over time, the sound will be perceived as noise, creating a “background,” referred to as the “Ground” condition. However, when several components become spectrally coherent over time, they form an auditory figure or object that perceptually “pops out” from the background, a condition referred to as the “Figure.”

**Figure 1:**
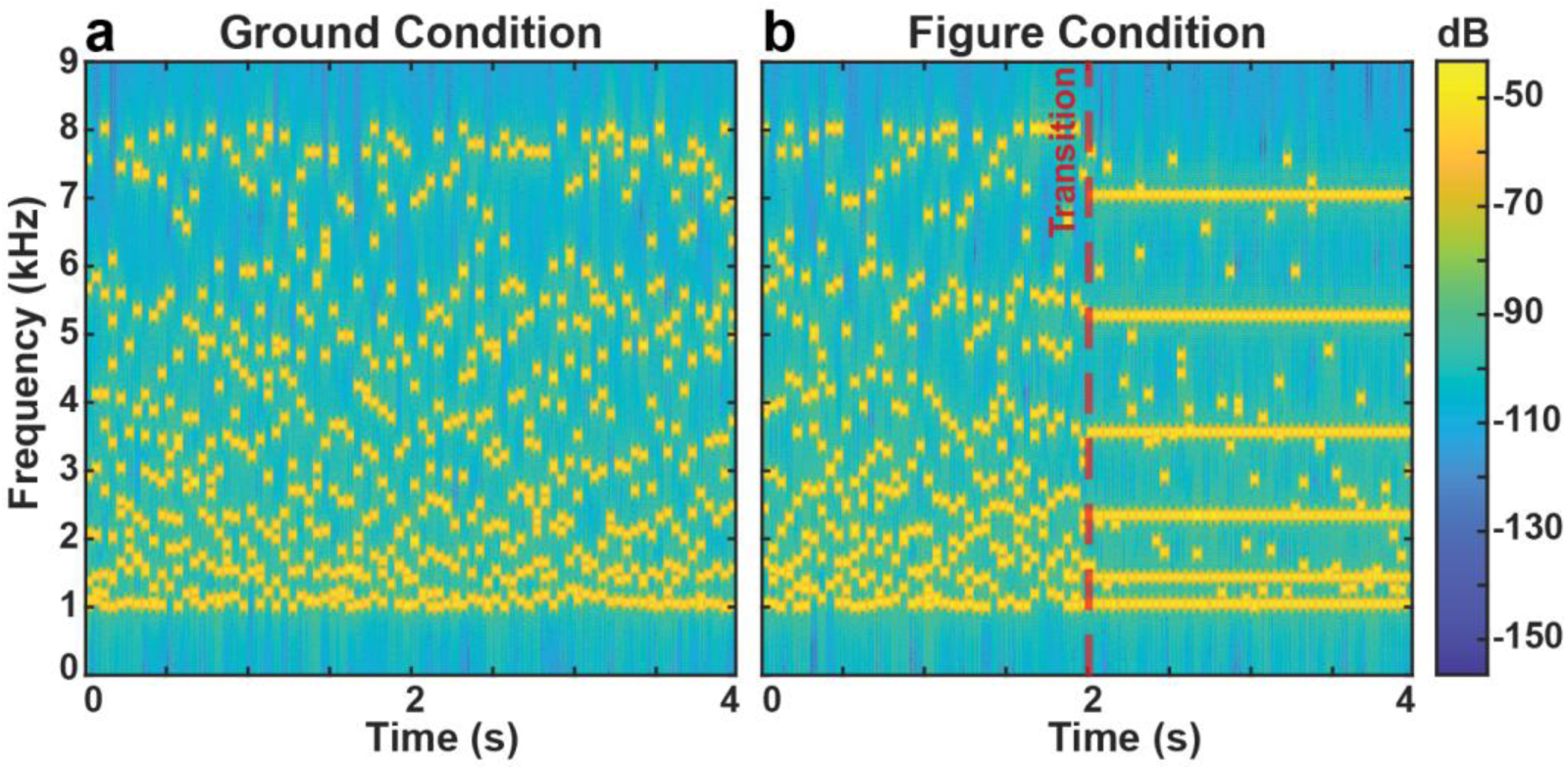
The spectrogram of the stochastic Figure-Ground (SFG) stimulus. The spectrograms of example stimuli used in the SFG paradigm for the Ground (panel **a**) and Figure (panel **b**) conditions are shown. In each of these conditions, the stimulus had a duration of four seconds. The Ground stimulus was composed of 40 segments, each with a duration of 50 ms. Each segment was composed of eight tones that were randomly selected from a pool of 145 components separated by 1/48^th^ of an octave in the frequency range of 1-8 kHz. The frequency distribution of the eight tones varied across segments. For the Figure stimulus, the first 20 segments were comprised of tones randomly distributed across frequency, identical to the Ground stimulus. The second 20 segments of the Figure stimulus included six tones with a fixed frequency across segments and two background tones with a varying frequency distribution across segments. These six coherent tones were 50 ms long and had at least a half-octave frequency separation within a segment. The red dashed line indicates the time when the transition to a “figure” occurs, which is fixed at the two-second mark. Note that tone pips fall between one and eight kHz, which is in the range of electric hearing provided by cochlear implant devices.

Neuroimaging studies using the SFG paradigm have shown that detecting auditory objects activates auditory regions in the brain, such as the superior temporal sulcus, medial geniculate body, and planum temporale (Teki et al., 2011), as well as non-auditory areas, including the inferior parietal sulcus, cingulate cortex, visual cortex, insula, and motor cortex (Holmes et al., 2021; Teki et al., 2011; Tóth et al., 2016a). The above studies highlight an extensive neural network contributing to auditory object detection, including auditory and non-auditory brain regions.

Studies using electroencephalography (EEG) and magnetoencephalography (MEG) have reported evoked responses elicited by SFG stimuli as a neural signature of auditory grouping. Passive listening to SFG stimuli elicited evoked responses that peaked ∼150 ms following the transition onset in the Figure condition, while evoked responses during active engagement in the SFG task showed a peak ∼200 ms after transition onset (O’Sullivan et al., 2015). This early evoked response can be interpreted as an automatic, pre-attentive neural process, which can be modulated by task attention (O’Sullivan et al., 2015). In the Figure condition, a low-frequency sustained “slow drift” has also been observed in the evoked responses (Guo et al., 2022; Teki et al., 2016), although no explanation for this response has yet been provided. In addition, using the SFG paradigm, two distinct event-related potentials were identified as a unique characterization of auditory figure-ground segregation (Boncz et al., 2024; Tóth et al., 2016a). The first is the object-related negativity (ORN), a negative deflection that emerges 200-300 ms post-transition onset (Boncz et al., 2024; Tóth et al., 2016a). The second is the P400 component that appears 450-600 ms following the emergence of an auditory object, which is elicited during active detection tasks (Tóth et al., 2016a). The ORN is believed to reflect the segregation of the “figure” from the background (Winkler & Denham, 2024), whereas the P400 is thought to reflect decision-making processes (Tóth et al., 2016a).

Outside of the SFG paradigm, studies on auditory stream segregation (also known as auditory streaming) have identified several brain oscillation responses in EEG. Low-frequency cortical activity in the delta (1-4 Hz) and theta (4-8 Hz) bands has been implicated in the segregation of multiple streams (Tóth et al., 2016b, 2020), the detection of a target sound embedded in background noise (McMullan et al., 2013; Ng et al., 2012), and selective attention to auditory targets (Plöchl et al., 2021; Viswanathan et al., 2019). Higher-frequency brain responses, reflected as modulations in alpha- and beta-band (8-30 Hz) oscillatory power, have been postulated to contribute to auditory streaming (de Vries et al., 2021; Müller & Weisz, 2012; Soltanparast et al., 2022; Wöstmann et al., 2019) and SiN perception more generally (Dimitrijevic et al., 2017; Price et al., 2019; Wöstmann et al., 2017). In particular, beta oscillations have been proposed to reflect predictions or expectations of future events (Arnal, 2012; Bastiaansen et al., 1999; Sedley et al., 2016; Todorovic et al., 2015), as a mechanism that might facilitate auditory object perception (Winkler et al., 2009). Alpha desynchronization is believed to play a role in the selective inhibition of a non-target stimulus in auditory (Strauß et al., 2014; Weisz et al., 2011) and visual perception (Feng et al., 2017; Foxe & Snyder, 2011).

In the presence of SNHL, brain responses to auditory stream segregation are modulated. Neural phase-locking to auditory stimuli in more general tasks is highly attenuated by SNHL (Buss et al., 2004; Mai & Howell, 2023; Woolf et al., 1981), and this degradation may be proportional to the degree of SNHL (Nash-Kille & Sharma, 2014). In particular, studies have shown attenuated phase-locking responses to SiN stimuli in CI users, which is manifested as a reduction in evoked response amplitudes in CI users compared to NH listeners (Burkhardt et al., 2022; Finke et al., 2016). It thus follows that phase-locking responses to auditory object detection may also be altered in listeners with SNHL. However, for SiN perception, the brain may employ mechanisms to compensate for degraded auditory inputs. For example, Price et al. (2019) reported increased beta-band connectivity between the prefrontal and auditory cortices with higher degrees of SNHL. Other studies have shown a modulation in alpha power during challenging SiN listening conditions in listeners with SNHL (Paul et al., 2021; Petersen et al., 2015). However, it is still unclear how SNHL affects delta and theta cortical processes underlying auditory stream segregation.

In principle, listeners who have SNHL but do not use a CI should be able to take advantage of their peripheral spectral and temporal resolution to a greater extent than CI users to detect auditory objects. This is due to the fact that in CI users, spectral and temporal resolution has been shown to be degraded, depending on CI electrode placement (Anderson et al., 2011; Bierer et al., 2015; Goehring et al., 2019) and the number of CI channels (Friesen et al., 2001; Henry & Turner, 2003). Decreasing electrode distance in the cochlea impairs auditory stream segregation (Chatterjee et al., 2006; Cooper & Roberts, 2007), and with fewer CI channels, the spectral resolution of sounds is further impoverished (Fu & Nogaki, 2005; Henry & Turner, 2003). These points imply that non-CI users with SNHL may have an advantage over CI users in spectral and temporal processing, provided that they are receiving sufficient amplification from their hearing aid (HA) devices. This idea has been supported by previous study findings showing enhanced spectral and temporal resolution in non-CI hard-of-hearing listeners (Henry et al., 2005) and HA users (Choi et al., 2018) relative to CI users.

Subjects with SNHL are able to detect auditory objects using the SFG paradigm. For example, Boncz et al. (2024) investigated the effect of mild hearing impairment on detecting auditory objects in the elderly. The authors reported that older people with SNHL needed a higher number of coherent tonal elements in the Figure condition to reach the same behavioral performance level compared to an age-matched NH population. In a study of 47 traditional CI users, 12% of the variability in SiN perception was explained by SFG performance, which was independent of peripheral spectral and temporal resolution measures (Choi et al., 2023). However, between these two previously tested populations—NH in Boncz et al. (2024) and traditional CI users in Choi et al. (2023)—there is a range of other listening configurations. For example, CI listeners who use both electric and acoustic hearing modalities ipsilaterally (i.e., hybrid CI users; Gantz & Turner, 2003) or contralaterally (i.e., bimodal CI users with CI stimulation on one side and HA on the other side; Illg et al., 2014) are said to have an electroacoustic (A+E) hearing configuration. Furthermore, listeners with traditional CIs that have long cochlear electrodes covering the cochlea from the base to the apex (Hochmair et al., 2015) rely on electric-only (E-only) stimulation for listening when they use such CI devices on both sides. E-only listeners are characterized by the lack of functional acoustic hearing, which can be defined as having unaided hearing thresholds below 80 dB (Gantz et al., 2022), on either ear. Thus, unlike the A+E group that receives acoustic stimulation from the non-CI ear combined with the electric stimulation from the CI ear, the E-only group receives solely electric stimulation from both implanted ears. Non-CI users, such as those who use HAs, have acoustic-only (A-only) auditory profiles, given that they receive bilateral acoustic stimulation. Critically, these groups of listeners may differ in the degree to which they experience spectral degradation, as CI users suffer from poorer spectral resolution than non-CI listeners with SNHL (Henry et al., 2005). Furthermore, among CI users, spectral resolution is degraded to a greater extent in listeners with E-only compared to A+E stimulation (Tejani & Brown, 2020). Nevertheless, how these different auditory configurations affect the detectability of auditory objects at the behavioral and neural levels is still not understood.

The role of cortical neural activity in auditory grouping and how it differs between listeners with and without SNHL remains largely unknown. Moreover, it is unclear whether behavioral and cortical measures of auditory object detection vary across different auditory configurations. The purpose of this study was to explore the role of cortical activity within the canonical frequency bands (delta, theta, alpha, and beta) in auditory object detection across different listening configurations. To this end, we implemented the SFG paradigm in the following groups: age-typical normal hearing (NH), acoustic-only (A-only), electroacoustic (A+E), and electric-only (E-only). We recorded EEG as the subjects performed the SFG task, to investigate the underlying brain mechanisms of auditory object detection in listeners with different hearing configurations. We hypothesized that listeners with SNHL would perform worse than NH listeners, that A-only listeners would perform better at the SFG task than CI users, and that E-only listeners would have poorer task performance compared to A+E listeners. Additionally, we hypothesized that phase-locking responses to the auditory objects would be degraded in listeners with SNHL, and to a greater extent in CI users compared to A-only listeners. For cortical activity measures, we sought to determine whether they contributed to task performance.

## 2. Methods

### 2.1 Participants

We recruited 85 participants who were divided into four groups: 20 NH listeners, 17 A-only bilateral HA users, 31 subjects with A+E CI configuration, and 17 subjects with E-only CI configuration. Subjects with SNHL were recruited from the University of Iowa Cochlear Implant Clinical Research Center, and non-CI users were recruited through email advertisements to the University of Iowa Community. Participants were excluded if they had severe neurological or cognitive deficits. All subjects were native speakers of American English, and all groups were matched by age. CI subjects needed at least six months of experience using the device to participate. Table 1 provides the demographic details of participants. This study was approved by the University of Iowa IRB ethics committee. Subjects signed a consent form and were reimbursed for participation.

**Table 1.** Demographics of the included participants.

|  | Groups |  |  |  | P value |
| --- | --- | --- | --- | --- | --- |
|  | NH<br>(N = 20) | A-only<br>(N = 17) | A+E<br>(N = 31) | E-only<br>(N = 17) |  |
| Age (yrs) | 58 (±10.87) | 66.53 (±13.6) | 63.81 (±11.79) | 58.71(±14.72) | 0.12 |
| Sex | M:8/F:12 | M:10/F:7 | M:15/F:16 | M:9/F:8 | - |
| Onset of HL (yrs) | N/A | 56.17 (±16.28) | 31.83 (±22.38) | 21.43 (±15.51)<sup>§</sup> | <0.001* |
| Age at HL intervention (yrs) | N/A | 58.41 (±16.12) | 42.81 (±19.5) | 25.25 (±16.25)<sup>§</sup> | <0.001* |
| Years of CI usage | N/A | N/A | 6.18 (±4.11) | 8.87 (±6.94)<sup>†</sup> | 0.401 |
| Unaided HFPTA | 19.13 (±9.48) | 57.72 (±16.92)<sup>‡</sup> | 93.53 (±14.51) | 106.43 (±8.25) | <0.001* |

\* Significant p-value obtained when testing with one-way ANOVA. <sup>§</sup>The results represent data for 16 subjects since data was missing from one subject. <sup>†</sup>Duration of CI usage was calculated by taking the average device usage for both ears. <sup>‡</sup> Unaided audiograms were missing for 3 subjects in this group, and therefore, the results represent the average of 14 subjects. HFPTA reflects the high-frequency pure tone average, which was measured by taking the average of hearing thresholds over the frequencies 1000 Hz, 2000 Hz, 4000 Hz, and 8000 Hz. HL = hearing loss. M = male, F = female. N/A = not applicable.

The NH group included listeners with age-typical hearing, which is characterized as mild to moderate sloping hearing loss at mid to high frequencies (1000 to 8000 Hz, Figure 2a). Listeners with SNHL were divided into either A-only, A+E, or E-only groups based on their device configuration. The A-only group included 17 bilateral HA users with (unaided) moderate to moderately severe SNHL in the high-frequency range (Figure 2b). The A+E group included a total of 31 subjects, all of whom had a CI on one side and a HA on the other side. Out of the 31 subjects, 8 were hybrid (listeners who received both electric and acoustic stimulation from the implanted ear, in addition to the acoustic stimulation from the unimplanted ear) and 23 were bimodal (listeners who received acoustic stimulation only from the unimplanted side while electric stimulation was received from the implanted side) CI users. The stimuli designed for this study were composed of tone pips ranging from 1000 to 8000 Hz in the frequency dimension (Figure 1), which is in the electric hearing range of all CI users. In other words, the presence of functional acoustic hearing in the low-frequency range (≤ 1000 Hz), as in the case of hybrid CI users, could not contribute to auditory object binding. Accordingly, hybrid and bimodal CI users did not principally differ in their auditory configuration for this task, and therefore, they were both included in the A+E group. All listeners in the A+E group had some degree of SNHL on the unimplanted ear, meaning that none of the listeners were considered to have single-sided deafness. Lastly, the E-only group included bilateral CI users with no functional acoustic hearing (i.e., 80 dB or less), meaning that E-only listeners had only electric stimulation on both sides.

**Figure 2.**
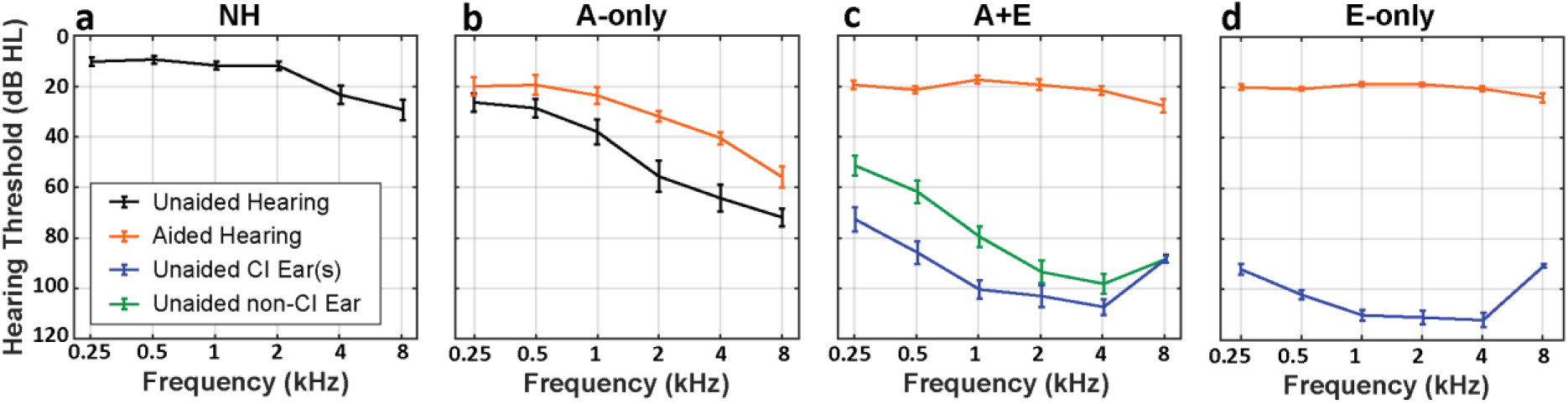
Aided and unaided audiometric results for the **a)** normal hearing (NH), **b)** acoustic-only (A-only), **c)** acoustic and electric (A+E), and **d)** electric-only (E-only) groups. For the NH, the audiogram depicts the average hearing thresholds over both ears. For the A-only group, aided (with the HA devices turned on) and unaided (with the HA devices turned off) hearing thresholds over both ears are shown. In the A+E group, the unaided (with the HA and CI devices turned off) audiograms show the average hearing thresholds over the CI and HA sides, separately. Aided audiograms depict hearing thresholds obtained for both ears with both the HA and CI devices turned on. For the E-only group, hearing thresholds averaged across ears are shown for aided (with both CI devices turned on) and unaided (with both CI devices turned off) conditions. The A-only group had 3 missing unaided audiograms and 4 missing aided audiograms. Error bars reflect S.E.M.

Conventional unaided audiometric testing was performed at the frequencies 250, 500, 1000, 2000, 4000, and 8000 Hz. In addition to the unaided audiometric thresholds, all listeners, except for the NH group, were tested in the aided condition. In the unaided conditions, participants were tested with pure tones using inserts or headphones, and in the aided condition, sound field testing was conducted using warble tones. Aided testing was conducted with both HA devices turned on for the A-only group, with the HA and CI devices turned on for the A+E group, and with both CI devices turned on for the E-only group. For most CI users, post-operative audiometric data were obtained from the date closest to the experiment time. However, in the absence of such data, hearing thresholds were substituted with the maximum frequency-specific intensity output of the audiometer (n = 11). For the A-only group, there were 3 and 4 missing audiograms for the unaided and aided conditions, respectively. Figure 2 illustrates audiometric results for all groups.

### 2.2 Stimuli

The SFG stimuli used in this study were the same as in Choi et al. (2023), which were developed following the same principles reported in Teki et al. (2011). We conducted extensive piloting with CI listeners to determine stimulus characteristics to allow for a range in performance across listeners. This led to a stimulus that consisted of 50-ms tone pip segments with eight frequency components selected at each time segment. The whole stimulus was 4 seconds long, and the time interval between each segment was 0 ms.

Two types of stimuli were used in this study: Figure and Ground. The “Ground” stimulus consisted of 40 segments. Each segment was composed of eight frequencies randomly selected from a distribution of 145 components separated by 1/48th of an octave across 1-8 kHz (Figure 1). The “Figure” stimulus was designed such that a transition from a ground to a figure always occurs at the 2-second mark. Like the Ground stimulus, the first half (20 segments) of the Figure stimulus consisted of eight tones per segment, randomly drawn from the same frequency pool. In contrast, the second half (20 segments) was constructed to include six frequencies that remained constant across time (forming a coherent auditory object). These six frequencies were selected such that each was at least half an octave apart, to avoid the potential spectral overlap in CI users with fewer than 10 channels. The other two components were randomly drawn from the remaining distribution for each segment.

The SFG paradigm contained 120 trials; half of these trials constituted the Figure condition, and the other half the Ground condition. In both conditions, tone pips were isolated to frequencies that would be in the electric hearing range of the CI listeners, i.e., from 1000 Hz to 8000 Hz. This was to ensure that the auditory signals could be encoded at the auditory periphery for all listeners.

### 2.3 Task

The experiment was carried out using a customized MATLAB script (The Mathworks, version R2022b) and Psychtoolbox 3 (Brainard & Vision, 1997). All subjects were naïve to the SFG stimuli. The experiment was carried out in a sound-controlled booth. SFG stimuli were delivered using a sound field loudspeaker (model LOFT40, JBL) positioned 1.2 m from the subjects at a zero-degree angle. Stimuli were presented at a sound level of 70 dB SPL. A screen was positioned approximately 1 meter in front of the subjects and was used to deliver visual instructions.

The presentation of the Figure and Ground trial conditions was randomized. Participants were asked to actively attend to the stimuli and determine whether they heard the emergence of the auditory object in the Figure trials by pressing on a keypad either “1,” indicating that they heard it, or “2,” indicating that they did not hear it. Subjects were instructed to respond after the sound offset and were given 5 seconds to respond on each trial. After each response, there was a delay period of 1 second, followed by a 1-second fixation cross (serving as the baseline), before the sound stimulus for the next trial was presented. A break was given every 40 trials. Throughout the experiment, participants were asked to keep their eyes fixated on a central cross and to minimize body and eye movements.

To test the hypothesis about task performance differences across the groups, we measured task accuracy, response bias, and reaction time. The accuracy of auditory object detection was measured as a d-prime value (d’), which is calculated by taking the difference between standardized hit rates for Figure trials and false alarm rates for Ground trials [d’ = z(Hit_Figure_) – z(FA_Ground_)]. In the case of extreme Hit_Figure_ and FA_Ground_ values, adjustment was applied as follows. A value of 0 was substituted with 0.5/i_n_, and a value of 1 was replaced with (i_n_-0.5/i_n_), wherein i_n_ is the number of trials recorded for that subject. To measure response bias, the criterion (c) was computed by taking the sum of the standardized hit rate for Figure trials and the false alarm rate for Ground trials, then multiplying the sum by −0.5 [c = −0.5*(z(Hit_Figure_) + z(FA_Ground_))]. For c measurement, extreme Hit_Figure_ and FA_Ground_ were corrected in a similar manner to the method described above. Reaction time (RT) was measured by taking the median RT across trials for a given subject.

### 2.4 EEG Acquisition

EEG data were recorded using the BioSemi active2 system with 64 channels, corresponding to the international 10/10 electrode placement system. EEG was sampled online at a rate of 2048 Hz. Two additional electrodes, the common mode sense (which serves as the ground electrode) and the driven right leg (which is used to reduce the amount of common-mode noise), were attached near the vertex. For CI users, electrodes that overlay the CI external components were removed prior to recording. Offset voltages were kept under 30 mV for all electrodes during the experiment.

### 2.5 EEG preprocessing

A customized MATLAB script was used to preprocess EEG data offline using EEGLAB toolbox functions (Delorme & Makeig, 2004). EEG data were downsampled by a factor of four to 512 Hz. The downsampled data were processed using a Butterworth bandpass filter (high-pass frequency: 2 Hz; low-pass frequency: 35 Hz; filter order: 3381). EEG recordings were epoched from −1000 ms to 5000 ms relative to the stimulus onset and then baseline corrected from −800 to −100 ms.

Removal of artifacts was done in a three-stage process, wherein the first stage involved removing outlier channels and trials, the second stage involved eliminating eye blinks and CI-related artifacts, and the third stage involved removing additional noisy trials following the removal of eye blinks and CI artifacts. For the first stage, a histogram of the maximum absolute voltage across all channels for that trial was plotted, whereby trials with extremely high voltages (about two standard deviations higher than the mean) were removed. Then, channels with maximum absolute voltages higher than 100 µV were excluded. Finally, the voltage of the remaining trials after excluding noisy channels was displayed as a histogram showing the maximum absolute voltage across all remaining channels for that trial, and bad trials were determined based on the same method described above. The number of remaining channels was 63.9 (± 0.31) in the NH, 63.71 (± 0.59) in the A-only, 57.72 (± 3.12) in the A+E, and 52.17 (± 4.79) in the E-only group.

For the second stage of the artifact removal process, we used independent component analysis (ICA) applying the Infomax algorithm (provided by the EEGLAB toolbox) to extract artifacts related to CI and eye and muscle movements. ICA components were determined as CI-related artifacts based on two criteria (Bakhos et al., 2012; Intartaglia et al., 2022). First, the presence of a strong direct current pedestal at the onset and offset of the sound stimulus. Second, the presence of a bipolar topography on the CI side. Muscle-related ICA artifacts were identified based on two criteria (Dharmaprani et al., 2016). First, the ICA time course shows continuous, large spikes, resembling electromyography waveforms. Second, topographical mapping of the ICA illustrates a single focal point of activity that has a peripheral head localization. This process resulted in removing an average of 1.95 (± 1.09) ICA components from the NH sample, 2.58 (± 1.12) from the A-only, 12.16 (± 3.79) from the A+E, and 11.11 (± 3.63) from the E-only groups.

In the third (last) stage of the artifact removal process, we rejected additional trials with excessive voltage in a similar manner as outlined above. Specifically, after the removal of ICA components, the maximum absolute voltage across all channels for each trial was calculated and trials with values higher than two standard deviations of the mean were excluded. This three-stage process resulted in an unequal number of trials between the Figure and Ground conditions. However, we equalized the number of trials to the condition that showed a smaller number of trials at the subject level, by taking the first equal count of trials. Subsequently, the total number of remaining trials for each of the Figure and Ground conditions was 49.25 (± 4.62) in the NH, 46.41 (± 5.25) in the A-only, 47.25 (± 5.38) in the A+E, and 40.47 (± 6.56) in the E-only group.

After cleaning the EEG data of artifacts, we applied the spherical spline interpolation method (Perrin et al., 1989) to restore channels that had been excluded either before the EEG recording or during the channel rejection phase. However, this interpolation method is known to be less reliable when applied to large clusters of missing channels (Perrin et al., 1989). This limitation is particularly relevant for CI users, in whom the temporoparietal channels are typically removed before EEG recording. To address this issue, we excluded 14 temporoparietal channels (seven on each side: C5, C6, CP5, CP6, T7, T8, TP7, TP8, P5, P6, P7, P8, P9, and P10) from all subsequent analyses. These channels were excluded in all groups to ensure comparability in the number of remaining channels.

### 2.6 Evoked responses

For evoked response analysis, we focused on five frontocentral channels (FZ, FCz, CZ, FC1, and FC2) based on prior evidence that EEG evoked responses to the SFG paradigm are largely driven by activity at these sites (Guo et al., 2022). After interpolation of missing channels, group-level evoked responses were measured by taking the average across the frontocentral electrodes, done separately for Figure and Ground trials. For subsequent processing of evoked responses, we re-referenced the EEG data to the average of O1 and O2 electrodes. These sites were chosen as the reference point for two reasons. First, these occipital areas are spatially distant from the CI device(s), which minimizes CI-related artifacts in the evoked responses (Intartaglia et al., 2022). Second, re-referencing to occipital sites provides a sufficient distance from frontocentral regions, enabling a reliable measurement of auditory N1 responses (Intartaglia et al., 2022). The re-referenced data were then filtered again with the same filter used in the preprocessing step, but with a low-pass frequency of 10 Hz and a filter order of 64.

Topographical maps of N1 responses were plotted for both sound onset and transition onset to determine whether the scalp activity pattern for auditory object detection was similar to that of the sound onset. For each subject, the N1 peak was identified as the maximum negative peak within two different time windows: 100-250 ms following the sound onset and 200-350 ms following the transition onset. These time frames were selected based on previous EEG findings using the SFG paradigm (Guo et al., 2022), showing N1 responses within these intervals. Subsequently, group-level N1 topographies were created by averaging N1 peaks across subjects within each group, separately for the sound and transition onsets. The 14 excluded temporoparietal channels (see section 2.5) were not included in the topographical mapping in any of the groups.

### 2.7 Time-frequency responses

Complex Morlet wavelets were used for time-frequency analysis as described in Cohen (2019). Morlet wavelets were created using a two-second time length centered at zero, with center frequencies extending in log_10_ space from 2 Hz to 30 Hz (20 frequencies). Each wavelet has a fixed 3-cycle width. The full width at half maximum was approximately equivalent to 1.126 seconds in the time domain and 0.785 Hz in the frequency domain. These Morlet wavelets were used for phase-locking and induced spectral power analyses, as detailed in the following subsections.

#### 2.7.1 Inter-trial phase coherence (ITPC)

The inter-trial phase coherence (ITPC) is a method used to measure the uniformity of phase angles and the alignment of phase-locked evoked responses. This analysis was performed to test our hypothesis about diminished phase-locking responses in the SNHL groups. Following Cohen (2014), the ITPC was calculated using the following formula:

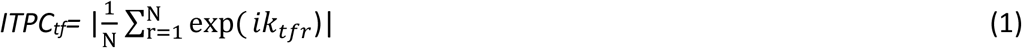

Where f is frequency, t is time, r is trial, and N is the number of trials. exp(ik) is taken from Euler’s formula, representing the polar distribution of the phase angle k over a given trial (r) for a time and frequency (tf) point. The ITPC was computed by convolving each trial with the Morlet wavelet at each frequency, from which the phase value was extracted from the resulting signal. ITPC values range from 0 to 1, reflecting the degree of consistency of phase coherence across trials, with 0 indicating no consistency and 1 indicating complete consistency.

The ITPC was quantified by averaging ITPC over trials in each subject for both the Figure and Ground conditions. Consistent with the evoked response analysis, ITPC was computed by taking the average across the frontocentral sites (FZ, FCz, CZ, FC1, and FC2), resulting in a two-dimensional ITPC dataset (time x frequency).

#### 2.7.2 Event-related spectral perturbations (ERSP)

To explore cortical activity underlying auditory object detection, we performed event-related spectral perturbation (ERSP) analysis. Morlet wavelets were applied to single-epoch EEG data to obtain time-frequency representations. Single-trial power was computed as the square magnitude of the complex signal. An initial baseline correction at the single-trial level was performed by converting power to decibels (dB) relative to the pre-stimulus baseline period (−800 to −100 ms), using the following formula:

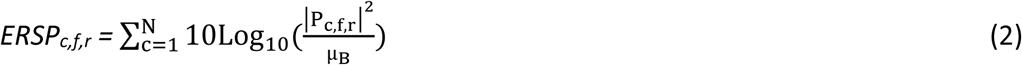

Whereby |P_c,f,i_|^2^ indicates the power of the single-trial EEG epochs and µ_*B*_ indicates the mean baseline power, measured by the formula:

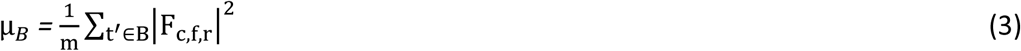

In both formulae (2 and 3), c is the channel, f is the frequency, N is the number of trials, and r is the trial. In the second formula, |F_c,f,i_|^2^ reflects the baseline power measured for each channel, frequency, and trial. *m* is the total number of time points in the baseline time interval, which was from −800 to −100 ms relative to sound onset. *B* is the individual time point in the baseline period. A relative power increase from the baseline is called an event-related synchronization (ERS), reflected as a positive ERSP value. Conversely, a relative power decrease from the baseline is known as an event-related desynchronization (ERD), reflected as a negative ERSP value.

For each subject, ERSP was averaged across trials, done separately for the Figure and Ground conditions. Subsequently, subject-level ERSP was averaged across the remaining 50 channels (see section 2.5). To facilitate valid between-group comparisons and account for inter-subject variability, each individual ERSP data were then normalized. This was achieved by z-scoring the data relative to the baseline period’s mean and standard deviation using the formula: Normalized ERSP = (ERSP – mean(baseline))/SD(baseline).

#### 2.7.3 Phase-locking and induced spectral power magnitude

We were interested in comparing the magnitude of induced spectral power and phase-locking activity related to auditory object detection across groups. For induced spectral power analyses, we extracted ERSP power in four frequency bands: delta (2–3.5 Hz), theta (4–7 Hz), alpha (8–15 Hz), and beta (17–30 Hz). These boundaries were chosen to align with the bin center frequency. For ITPC analyses, the amount of phase-locking value was quantified in the delta and theta bands combined, based on previous studies that showed phase-locking responses to auditory streaming in these frequency bands exclusively (Ng et al., 2012; Plöchl et al., 2022; Riecke et al., 2015).

A cluster-based permutation approach (Maris & Oostenveld, 2007) was applied to ERSP and ITPC data to detect significant clusters for each frequency band in each group. The cluster-based permutation test was performed using two-sided paired t-tests on the group-level ERSP and ITPC data between Figure and Ground conditions from 2 to 4 seconds, the time of auditory object emergence in the Figure condition. For each frequency band, we took the average across frequency bins that fall within the frequency band range. As a result, the cluster permutation test was carried out on two-dimensional data (time x subject) for each frequency band separately, at the alpha level of 0.05 and with 5000 permutations. The minimal cluster size was set to be at least 50 ms, and clusters were determined to be significant if *p*-values were less than 0.05. In cases where no significant clusters were found in a particular frequency band for a given group, the magnitude of ERSP or ITPC was not computed.

To visualize the scalp distribution of frequency-specific ERSP and ITPC measures, topographical maps were plotted, reflecting averaged responses within the significant clusters. Like N1 topographies, the scalp mapping for ERSP and ITPC measures were shown for the 50 remaining channels, i.e., the 14 excluded temporoparietal electrodes were not included in these topographies. No topographies were plotted for the group(s) that showed no significant clusters.

### 2.8 Statistical Analyses

Statistical analyses were performed in MATLAB (version R2023b), SPSS (version 29), and R (version 4.3.1). To determine whether the main effect of task performance (d’, c, and RT), ERSP spectral power (delta, theta, alpha, and beta), and ITPC magnitude differed across groups, we performed one-way ANOVAs. If fewer than three groups had significant clusters for a given frequency-specific ERSP or ITPC magnitudes (detected using the cluster-based permutation approach; see section 2.7.3), we used independent-sample t-tests to compare these effects between the remaining two groups. Planned post-hoc analyses using three orthogonal contrasts were carried out whenever omnibus ANOVA results were significant, as follows:

- Contrast 1 (NHvSNHL): This contrast compared the NH and SNHL groups. The SNHL group was composed of the A-only, A+E, and E-only groups. The contrast coefficient was −1 for the NH group and 0.33 for each of the A-only, A+E, and E-only groups. This contrast allowed us to ask whether listeners with SNHL, in general, differ from NH listeners on the basis of the measures described above.
- Contrast 2 (HAvCI): This contrast compared measures between HA and CI users. Here, CI users include both the A+E and E-only groups. The contrast coefficient was set to 0 for the NH, 1 for the A-only, and −0.5 for the A+E and E-only groups. This contrast enabled us to ask whether cochlear implantation, in particular, affects the task performance and neural measures that differ from listeners with SNHL who do not use a CI.
- Contrast 3 (AEvE): This contrast focused on CI users, comparing measures between the A+E and E-only groups, which tested whether functional acoustic hearing influenced outcomes. The contrast coefficient was 0 for both the NH and A-only groups, 1 for the A+E group, and −1 for the E-only group.

These contrasts were constructed hierarchically, such that each subsequent contrast evaluated a more specific distinction among auditory profiles, enabling an examination of how different listening configurations impact behavioral and cortical measures of auditory object detection. As a final step, we performed a hierarchical multiple regression analysis that aimed to determine the unique contribution of listening configurations and cortical responses to SFG task performance. Hierarchical regression models are designed to isolate the contribution of each variable, even when collinearity exists among predictors (McMurray et al., 2025).

For all statistical analyses, results were considered significant when the *p*-value was less than 0.05. Levene’s test was performed to evaluate whether the variance is equal across the groups for each of the tested measures. In the case that the assumption of equal variance is violated, Welch’s ANOVA and Welch’s t-test were performed instead. Effect size results for ANOVA (eta squared, *η*^2^) and contrast (Cohen’s d) testing were reported as appropriate.

## 3. Results

### 3.1 Task performance

Levene’s test revealed an unequal variance for d’ (F_3,81_= 7.138, *p* < 0.001) and RT (F_3,81_= 2.845, *p* = 0.043), but not for c (F_3,81_= 1.188, *p* = 0.319). Task accuracy results are illustrated in Figure 3a, demonstrating high sensitivity to the auditory object in the NH and A-only listeners compared to A+E and E-only listeners; however, all groups performed above chance level (NH: mean = 4.386, SD = 0.466; A-only: mean = 3.822, SD = 1.127; A+E: mean = 2.566, SD = 1.412; E-only: mean = 2.013, SD = 1.442). A one-way Welch’s ANOVA on d’ showed that auditory object detection was significantly different between groups (F_3,36.576_= 25.102 *p* < 0.001, *η*^2^ = 0.365). Post-hoc analysis revealed that NH participants had higher d’ than SNHL (NHvSNHL: t_66.370_ = −7.739, *p* < 0.001, Cohen’s d = −1.292) and that HA users were more accurate at detecting auditory objects than CI users (HAvCI: t_31.769_ = 4.067, *p* < 0.001, Cohen’s d = 1.242). However, performance accuracy did not differ between the A+E and E-only CI users (AEvE: t_32.476_ = 1.280, *p* = 0.210, Cohen’s d = 0.448).

**Figure 3.**
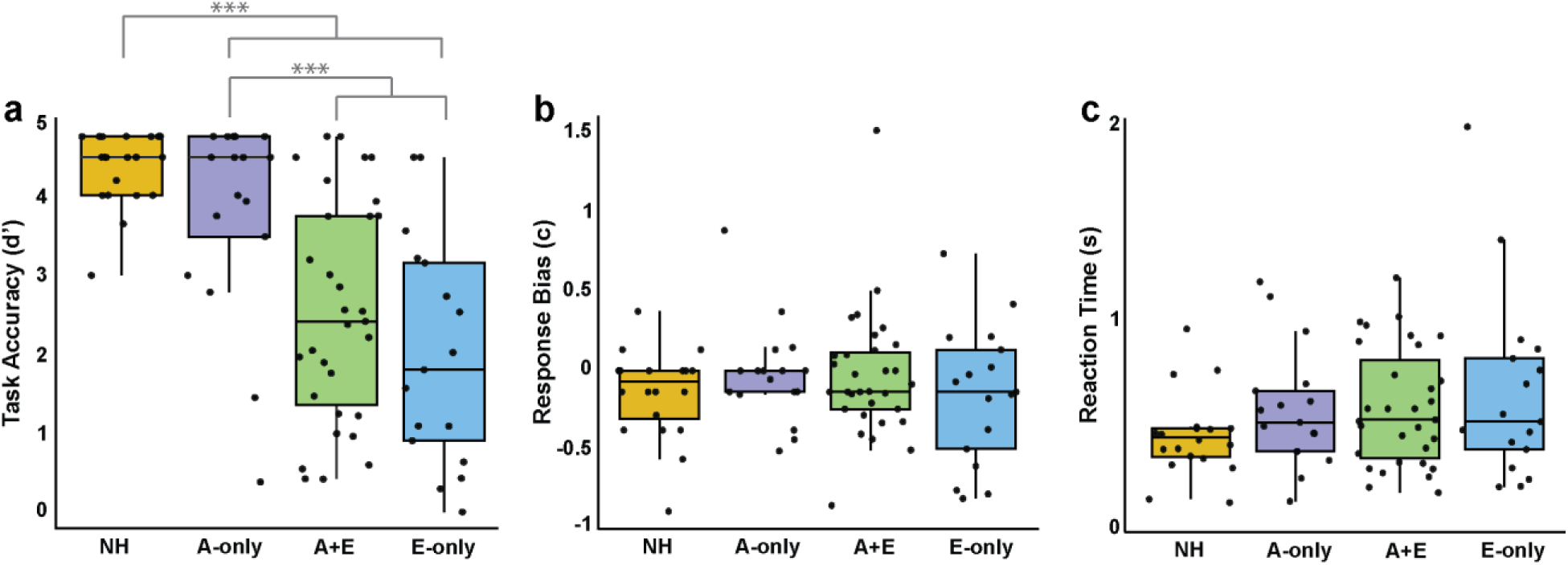
SFG task performance. Panel **a** shows auditory object detection accuracy (d’), panel **b** shows response bias (c), and panel **c** shows the reaction time results. Dots refer to individual data points. *** p < 0.001.

The response bias (c) was below zero for all groups (NH: mean = −0.136, SD = 0.277; A-only: mean = −0.022, SD = 0.317; A+E: mean = −0.031, SD = 0.401; E-only: mean = −0.078, SD = 0.366), indicating a slight tendency to respond as “1” (i.e., that the auditory object was detected, even when it was not present). The results for response bias are depicted in Figure 3b. One-way ANOVA showed that c did not significantly differ across groups (F_3,81_= 0.703, *p* = 0.553, *η*^2^ = 0.025).

The results for RT showed a different pattern (Figure 3c), with the fastest responses seen among NH listeners (NH: mean = 0.481, SD = 0.194; A-only: mean = 0.593, SD = 0.275; A+E: mean = 0.601, SD = 0.270; E-only: mean = 0.670, SD = 0.433). However, this pattern was not statistically significant, as indicated by one-way Welch’s ANOVA results showing no difference in RT between groups (F_3,38.791_= 1.704, *p* = 0.182, *η*^2^ = 0.046). Overall, these results indicate that the accuracy of detecting auditory objects is diminished in listeners with SNHL, particularly in those with CI devices.

### 3.2 Evoked responses

Evoked responses were measured at the frontocentral channels for both the Figure and Ground conditions. Figure 4a illustrates group-level evoked responses at frontocentral sites for both conditions. We observed three evoked responses: at sound onset, transition onset, and sound offset. Our analyses only focused on the evoked responses following the sound and transition onsets.

**Figure 4.**
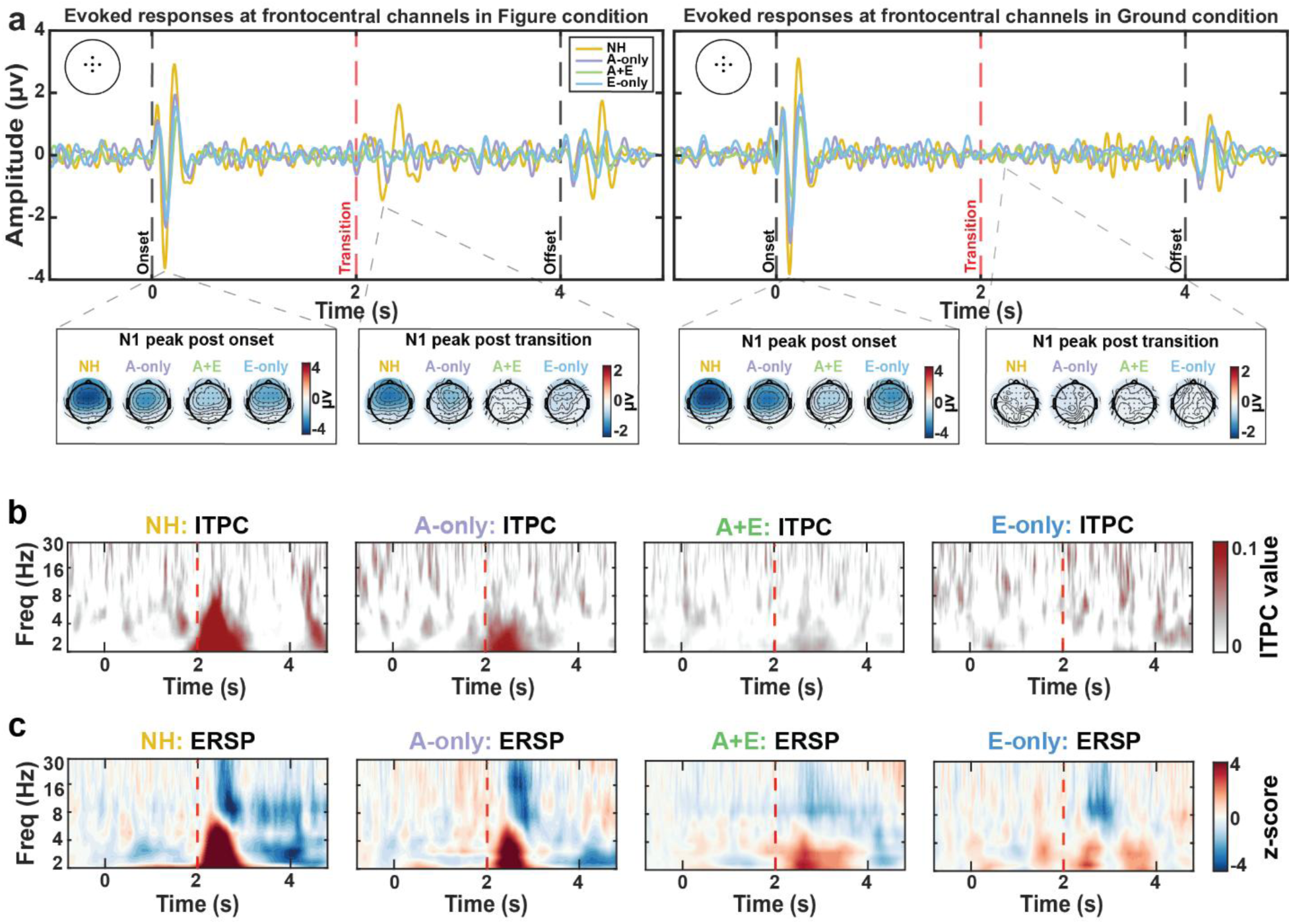
EEG results of the SFG paradigm. Panel **a** shows the evoked responses measured at the frontocentral sites (FZ, FCz, CZ, FC1, and FC2) for both the Figure and Ground conditions in NH (yellow), A-only (purple), A+E (green), and E-only (blue) groups. The topographical distribution of evoked responses represents the N1 peak measured within a 150-ms time window following the sound (from 0.10 to 0.25 seconds) and transition (from 2.20 to 2.35 seconds) onsets. Note that topographical mapping for 14 temporal sites (seven on each side) is excluded for all groups (see methods). Panel **b** depicts intertrial phase coherence (ITPC) results for each group. ITPC was measured across frontocentral channels by subtracting Ground from Figure ITPC. Panel **c** illustrates group-level ERSP measured across all 50 remaining electrodes by taking the difference between the Figure and Ground conditions. Positive ERSP values reflect event-related synchronization activity, and negative ERSP values indicate event-related desynchronization activity. Vertical dashed red lines show the transition onset, i.e., the time of auditory object emergence.

For all groups, the evoked responses following the sound onset in both conditions had a P1-N1-P2-N2 complex, wherein the N1 component peaked at ∼130 ms in the NH group. The topography of this N1 peak showed a fronto-central distribution, which looked similar between the Figure and Ground conditions. Relative to the NH group, weaker frontocentral activity for Figure and Ground conditions was shown in the other groups.

For the transition onset, the evoked response in the Figure condition appeared as a P1-N1-P2-N2 complex in the NH group, where the N1 component of this complex showed a peak at ∼264 ms relative to the transition onset. Strikingly, the A+E and E-only groups did not show any visible evoked responses following the transition onset in the Figure condition; however, relative to the NH group, a weaker evoked response appeared in the A-only group. For the NH group, the topography of the transition-elicited N1 peak (in the Figure condition) revealed a fronto-central pattern, similar to the sound-onset elicited N1 peak topography. The topography of the other groups was dissimilar to that of the NH group, showing no clear pattern. In the Ground condition, the topography map at the time window of N1 peak for the Figure condition showed no localization of activity for all groups, consistent with the absence of evoked responses in this condition. These results suggest that the neural generators of auditory object-related evoked responses are similar to those elicited by the sound onset, which were diminished in individuals with SNHL.

### 3.3 Time-frequency responses

The next analysis addressed two aims. First, we sought to identify the period during which power was significantly different between the Figure and Ground conditions. Second, we asked whether EEG measures in the Ground condition (i.e., Ground-only stimulus) differed across the groups. For EEG analyses, we focused on five outcomes: ITPC, delta-, theta-, alpha-, and beta-ERSP.

For the first aim, we performed cluster-based permutation testing at a group level, comparing ITPC and frequency-specific ERSP responses between the Figure and Ground conditions. This method allowed us to find the time for each group at which the magnitude of ITPC and ERSP differed between conditions. The results are provided in Table 2, showing one cluster per measure in the NH group. No significant clusters were found for ITPC in the A+E and E-only groups. For theta and delta ERSP, among SNHL groups, only the A+E group showed a single cluster for delta ERSP. However, all SNHL groups exhibited one cluster for alpha and beta ERSP.

**Table 2.** The results of cluster-based permutation tests showing the timing of significant clusters in ERSP and ITPC data for each group.

|  | NH |  |  |  | A-only |  |  |  | A+E |  |  |  | E-only |  |  |  |
| --- | --- | --- | --- | --- | --- | --- | --- | --- | --- | --- | --- | --- | --- | --- | --- | --- |
|  | Time (sec) |  | t value | p value | Time (sec) |  | t value | p value | Time (sec) |  | t value | p value | Time (sec) |  | t value | p value |
|  | start | end |  |  | start | end |  |  | start | end |  |  | start | end |  |  |
| ITPC | 2.001 | 3.037 | 2.72E+03 | <0.001 | 2.113 | 2.679 | 819.195 | 0.022 | N/C |  |  |  | N/C |  |  |  |
| Delta | 2.041 | 2.873 | 2.00E+03 | 0.005 | N/C |  |  |  | 2.238 | 3.651 | 2.41E+03 | 0.006 | N/C |  |  |  |
| Theta | 2.167 | 2.605 | 895.205 | 0.021 | N/C |  |  |  | N/C |  |  |  | N/C |  |  |  |
| Alpha | 2.386 | 2.933 | -1032.22 | 0.011 | 2.470 | 3.003 | -967.869 | 0.009 | 2.490 | 3.490 | -1.67E+03 | 0.0026 | 2.677 | 3.146 | -702.327 | 0.018 |
| Beta | 2.355 | 2.792 | -875.921 | 0.001 | 3.466 | 3.992 | -872.214 | 0.010 | 2.642 | 2.865 | -403.344 | 0.019 | 2.802 | 2.976 | -305.197 | 0.033 |

N/C means no significant clusters were found by the Monte Carlo permutation test.

For the second aim, we asked whether the Ground condition differed across the groups for each of the five outcomes. This step was performed before subtracting the Ground condition from the Figure condition to rule out that any potential differences across the groups are not driven by differences in the Ground condition. Given that the Ground condition serves as a control condition in our experiment, we did not expect differences across the groups concerning any of the outcomes. For frequency-specific ERSP and ITPC outcomes, the magnitude of ERSP and ITPC measures was calculated by averaging responses within the significant clusters revealed by the cluster-based permutation tests (Table 2). Since significant clusters only existed for the NH and A-only groups for ITPC and NH and A+E groups for delta ERSP, an independent sample t-test was performed, as Levene’s test showed equal variance between the groups for both measures (*p* > 0.05). The results revealed no significant differences (*p* > 0.05) across the groups for the ITPC and delta ERSP measures. For alpha and beta ERSP, one-way ANOVA was performed to examine the difference in Ground condition across all groups after confirming that the variance was equal (Levene’s test showed a p-value greater than 0.05). The results showed that only beta ERSP was significantly different across groups (F_3,81_ = 2.862, *p* = 0.041, *η*^2^ = 0.096).

Since the Ground condition differed across the groups only in one outcome (beta ERSP), our subsequent EEG analyses used the difference between Figure and Ground conditions. That is, the magnitude of each of the five measures was computed by averaging values across time within the significant clusters, where identified, from the Figure minus Ground contrast. A depiction of the ITPC and ERSP results for each of the Figure and Ground conditions is provided in Supplementary 1 and 2, respectively. The following subsections present detailed results for each of the five measures.

#### 3.3.1 ITPC

The NH and A-only groups showed an increase in ITPC responses following the transition onset, whereas no such responses were seen in the A+E and E-only groups, as the cluster-based permutation test revealed a lack of clusters in these two groups (Table 2). ITPC appeared to be the strongest in the NH group, where the increase in ITPC value was noted in the delta and theta bands. Figure 4b depicts the ITPC results for each group.

The magnitude of ITPC responses is illustrated in Figure 5a. Levene’s test showed the variance was not equal between the NH and A-only groups (F_1_,_35_= 4.236, *p* = 0.047). Comparing the magnitude of ITPC responses between the NH (mean = 0.094, SD = 0.075) and A-only (mean = 0.036, SD = 0.048) groups, Welch’s independent-sample t-test revealed a significant difference (t = 3.060*, p* = 0.004). The topography of the ITPC response in the NH group showed a fronto-central distribution, which appeared to a weaker extent in the A-only group (Figure 6a). These findings support the observed lack of evoked responses to auditory object emergence in CI listeners

**Figure 5.**
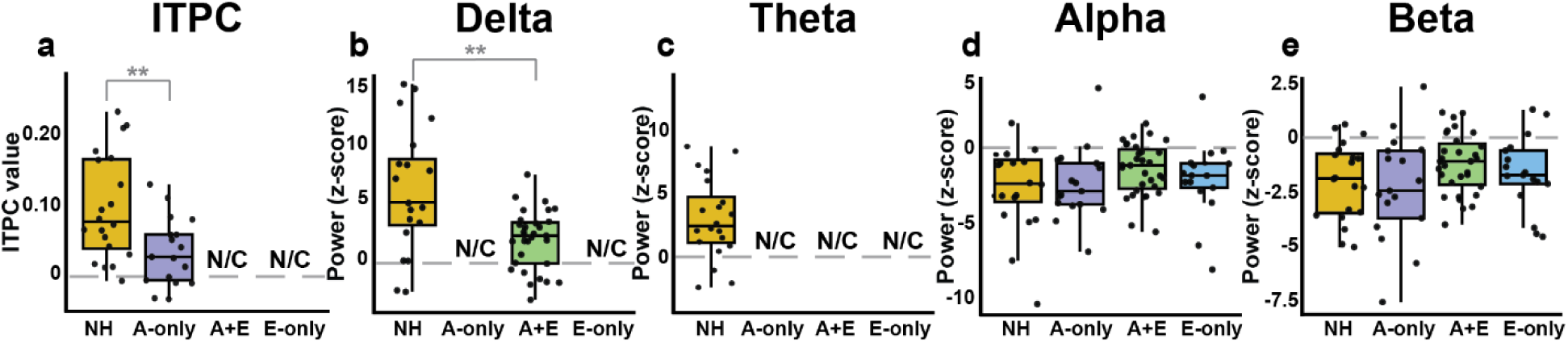
The difference in the magnitude of phase-locking and induced spectral activity across groups. For panels **a**-**e**, the magnitude was calculated from within the significant clusters in the Figure minus Ground ERSP and ITPC for each group. ITPC power in each group is illustrated in panel **a**. The group-level magnitude of the delta (2 – 3.5 Hz), theta (4 –7 Hz), alpha (8–15 Hz), and beta (17–30 Hz) ERSP power is demonstrated in panels **b, c**, **d**, and **e**, respectively. Individual data points are shown as dots. N/C means no significant clusters were found by the Monte Carlo permutation test. ** p < 0.01.

**Figure 6.**
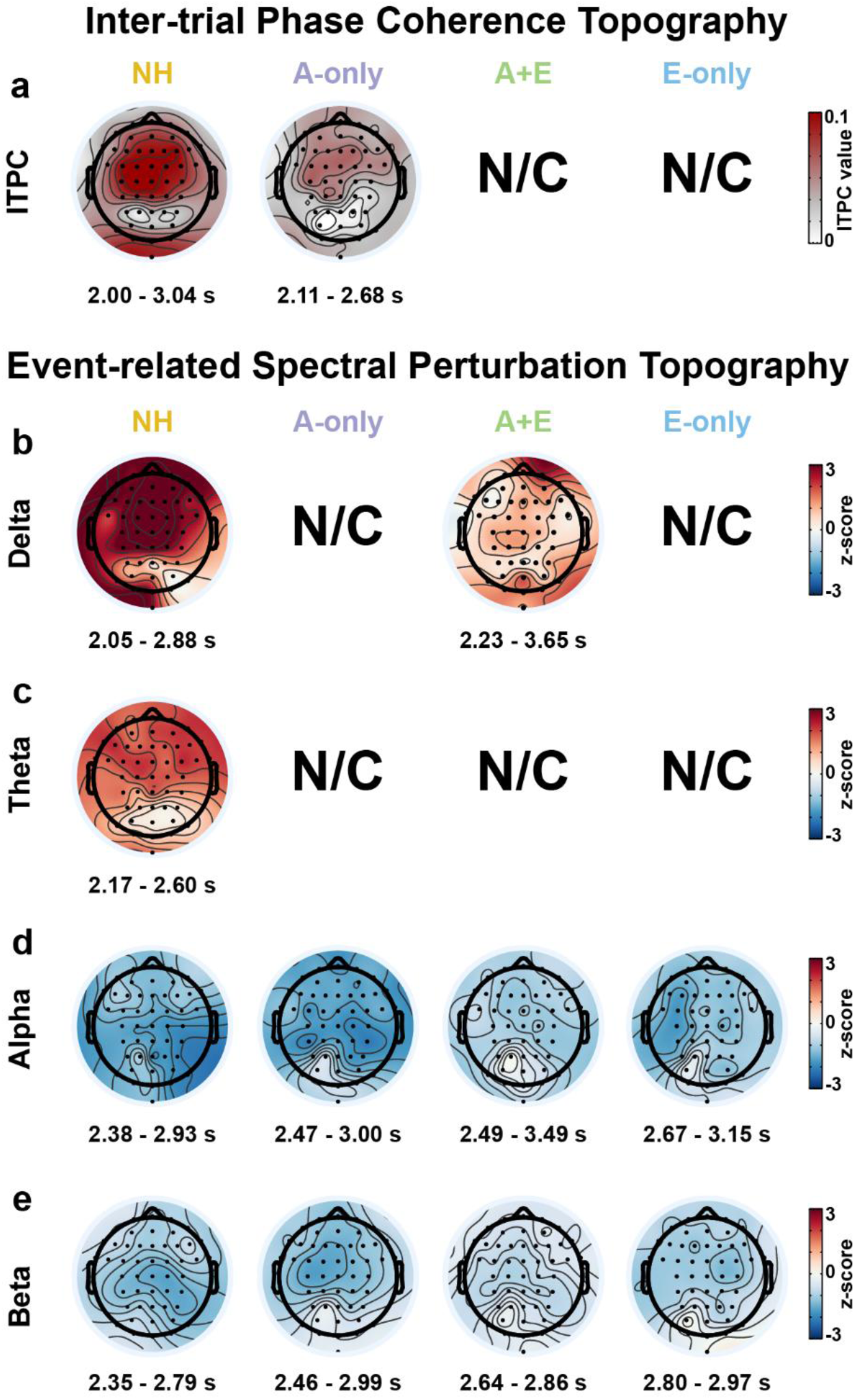
The topography of intertrial phase coherence (ITPC) and event-related spectral perturbation (ERSP) activity. Topographies represent the ITPC and frequency-specific ERSP magnitude measured within the significant cluster timing. Panel **a** represents the topography for ITPC responses. Panels **b**, **c**, **d**, and **e** show the topography of delta (2 – 3.5 Hz), theta (4 –7 Hz), alpha (8–15 Hz), and beta (17–30 Hz) ERSP activity, respectively. Note that topographical mapping for 14 temporal sites (seven on each side) is excluded for all groups (see methods). The start and end points of significant clusters are shown below the topographies. N/C means no significant clusters were found by the Monte Carlo permutation test.

#### 3.3.2 Delta ERSP

An increase in delta synchronization (ERS) was observed in the NH and A+E groups following the transition onset. Figure 4c illustrates ERSP results for each of the groups. The cluster-based permutation test showed a significant cluster only for the NH and A+E groups (Table 2).

The magnitude of delta ERS for the NH (mean = 6.146, SD = 5.302) and A+E (mean = 1.966, SD = 2.515) groups is depicted in Figure 5b. Levene’s test showed that the NH and A+E groups had unequal variance (F_1_,_49_= 13.308, *p* < 0.001). The magnitude of delta ERS was significantly higher in the NH compared to the A+E group (Welch’s independent-sample t-test: t_49_= 3.294, *p* = 0.003). The topography of delta ERS responses is depicted in Figure 6b, showing a clear frontocentral activity in the NH group. A similar topographical mapping was observed in the A+E group, albeit being weaker than the NH group. Notably, the delta ERS topography resembled the ITPC topography in the NH group, showing a similar frontal and central scalp distribution, suggesting that delta power reflects stimulus-driven phase-locking responses to the emergence of the auditory object.

#### 3.3.3 Theta ERSP

Similar to delta ERS, an increase in theta power was also observed following the transition onset. As mentioned previously, Monte Carlo cluster permutation tests revealed significant clusters only for the NH group, indicating that listeners with SNHL do not have theta ERS elicited by the emergence of the auditory object.

The magnitude of theta power for the NH group (mean = 3.149, SD = 3.150) is shown in Figure 5c. Topographical representation of theta ERS also shows a frontocentral scalp distribution in the NH group, as seen in Figure 6c. Along with delta ERS, theta topographical mapping suggests that theta might also be reflecting phase-locking responses to the auditory figure emergence, suggested by its similarity to ITPC topography.

#### 3.3.4 Alpha ERSP

A decrease in alpha power (ERD) emerged after the transition onset in all groups. All groups had significant clusters obtained from the Monte Carlo permutation test (Table 2), confirming that alpha ERD activity to auditory object emergence was present in all groups.

Group-level alpha ERD magnitude is depicted in Figure 5d, showing all groups have similar levels of alpha power (NH: mean = −2.552, SD = 2.768; A-only: mean = −2.269, SD = 2.373; A+E: mean = −1.319, SD = 1.744; E-only: mean = −1.946, SD = 2.478). All groups had equal variance, as indicated by Levene’s test (F_3,81_= 0.760, *p* = 0.520). One-way ANOVA showed that alpha power was not significantly different across groups (F_3,81_= 1.350, *p* = 0.264, *η*^2^ = 0.048). These results indicate that alpha induced spectral responses are present in all groups, which are not affected by peripheral auditory configuration. Figure 6d shows alpha ERD topography for each group. Across all groups, the alpha ERSP topography showed a wide distribution of scalp activity encompassing frontal, central, and parietal regions.

#### 3.3.5 Beta ERSP

Beta ERD appeared in all groups, showing a similar trend to alpha ERD. Monte Carlo permutation tests showed that all groups exhibited significant clusters (Table 2), which confirms the presence of strong beta ERD in all groups.

The magnitude of beta ERD is illustrated in Figure 5e, showing comparable levels of beta ERD magnitude across the groups (NH: mean = −1.925, SD = 1.767; A-only: mean = −2.235, SD = 2.441; A+E: mean = −1.140, SD = 1.373; E-only: mean = −1.568, SD = 1.708). Levene’s test showed that the variance was equal across groups (F_3,81_= 2.092, *p* = 0.545). One-way ANOVA performed on beta ERD found no significant differences across the groups (F_3,81_= 1.618, *p* = 0.192, *η*^2^ = 0.057). The topography of beta ERSP power showed localized activity in the central and parietal areas of the NH and A+E groups (Figure 6e). In contrast, this activity was more centrally localized in the remaining groups. Overall, the findings suggest that, similar to alpha ERD, beta ERD is not influenced by the auditory profile of the listeners.

### 3.4 Predictors of SFG performance

We have shown that task accuracy (d’) was different between groups and that certain EEG measures (i.e., delta and theta ERSP power) were not present in all listeners with SNHL. Additionally, we found that ITPC responses were present in the NH and A-only groups, but not in CI users. However, all groups exhibited alpha and beta activity following the auditory object emergence, and they were not significantly different between contrasts. The pattern of findings suggests that low-frequency (delta and theta) ERSP responses are degraded in listeners with SNHL, and that phase-locking (ITPC) responses are diminished in CI users, implying that such responses are likely to be driven by acoustic hearing. However, alpha and beta ERD did not show statistically significant differences between any contrasts, suggesting that these measures index event-related cortical activity that is not affected by the degree of SNHL or auditory profile. Table 3 summarizes the overall results obtained from the planned post-hoc contrast analyses.

**Table 3.** Summary of planned contrast analyses’ results.

| Measure | NHvSNHL contrast |  |  | HAvCI contrast |  |  | AEvE contrast |  |  |
| --- | --- | --- | --- | --- | --- | --- | --- | --- | --- |
|  | Cohen's d | t value | p value | Cohen's d | t value | p value | Cohen's d | t value | p value |
| $d'$ | **-1.292** | **-7.739** | **<0.001** | **1.242** | **4.067** | **<0.001** | 0.448 | 1.280 | 0.210 |
| Alpha | 0.308 | 1.194 | 0.236 | -0.278 | -0.972 | 0.334 | 0.274 | -0.906 | 0.367 |
| Beta | 0.156 | 0.906 | 0.548 | -0.494 | -1.728 | 0.088 | 0.240 | 0.794 | 0.430 |

Accordingly, we ran a hierarchical regression to examine whether alpha and beta ERSP contribute to auditory object detectability across different auditory configurations. The model specifically aimed to assess the contribution of cortical activity that was present in all groups to a comparable level. This approach enabled the investigation of the role of neural activity that is common across listeners with different auditory configurations in task accuracy. The first level of the model included the three planned contrasts to determine whether hearing configuration predicts task performance. In the second level of the model, we added alpha and beta ERD to determine the contribution of cortical processes elicited during auditory object processing (indexed by alpha and beta induced spectral power) to SFG task performance (d’), beyond peripheral hearing profiles.

In the first level of the hierarchical regression model, we included the three hearing profile contrasts as predictors of SFG d’ (4).

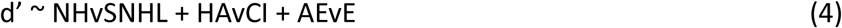

The first level model (4) was significant (F_3,81_ = 15.5, *p* < 0.001, adjusted *R^2^* = 0.341). In this model, both the NHvSNHL and HAvCI contrasts significantly predicted d’ (both *p* < 0.001). A full summary of Level 1 results is provided in Table 4. The results from Level 1 suggest that SFG performance is predicted by whether listeners have SNHL, and if so, whether individuals use a CI.

**Table 4.** Hierarchical multiple regression model analysis results (N = 85)

| Level 1: $d' \sim \text{NHvSNHL} + \text{HAvCI} + \text{AEvE}$ | | | | |
| --- | --- | --- | --- | --- |
| $d'$ | $\beta$ | SE | t value | p value |
| Intercept | 0.038 | 0.091 | 0.427 | 0.670 |
| NHvSNHL | **-0.785** | **0.157** | **-4.980** | **< 0.001** |
| HAvCI | **0.672** | **0.154** | **4.349** | **< 0.001** |
| AEvE | 0.182 | 0.122 | 1.486 | 0.141 |
| Level 2: $d' \sim \text{NHvSNHL} + \text{HAvCI} + \text{AEvE} + \text{Alpha} + \text{Beta}$ | | | | |
| $d'$ | $\beta$ | SE | t value | p value |
| Intercept | 0.027 | 0.088 | 0.307 | 0.759 |
| NHvSNHL | **-0.744** | **0.154** | **-4.824** | **< 0.001** |
| HAvCI | **0.602** | **0.153** | **3.944** | **< 0.001** |
| AEvE | 0.214 | 0.119 | 1.791 | 0.077 |
| Alpha | -0.094 | 0.116 | -0.809 | 0.421 |
| Beta | -0.161 | 0.117 | -1.371 | 0.174 |

In level 2 of the model, alpha and beta were added to explore whether cortical processes additionally contribute to d’ (5).

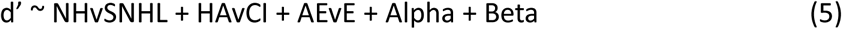

The addition of alpha and beta to the model significantly accounted for an additional 4% of the variance (F_2,79_ = 3.524, *p* = 0.034). A summary of the Level 2 model is provided in Table 4. The Level 2 model was also significant (F_5,79_ = 11.29, *p* < 0.001, adjusted *R^2^* = 0.379), with significant NHvSNHL and HAvCI contrasts (*p* < 0.001). Importantly, neither alpha nor beta was a statistically significant predictor in level 2 (*p* > 0.05). However, the fact that the addition of both was significant overall and the fact that these were correlated with each other (r = 0.671, *p* < 0.001) raises the possibility of a suppression effect where shared variance between alpha, beta, and d’ prevents either from becoming individually significant.

To test whether alpha or beta had a suppressive effect in the Level 2 model, we performed post-hoc analyses where either alpha or beta was removed from the Level 2 (5) model. The results showed that alpha (t_80_ = −2.261, *p* = 0.026) and beta (t_80_ = −2.534, *p* = 0.013) were statistically significant when either variable was added separately into the level 1 model (Figure 7). Collectively, these findings suggest that alpha and beta oscillations may reflect a common neural mechanism for auditory object detection.

**Figure 7.**
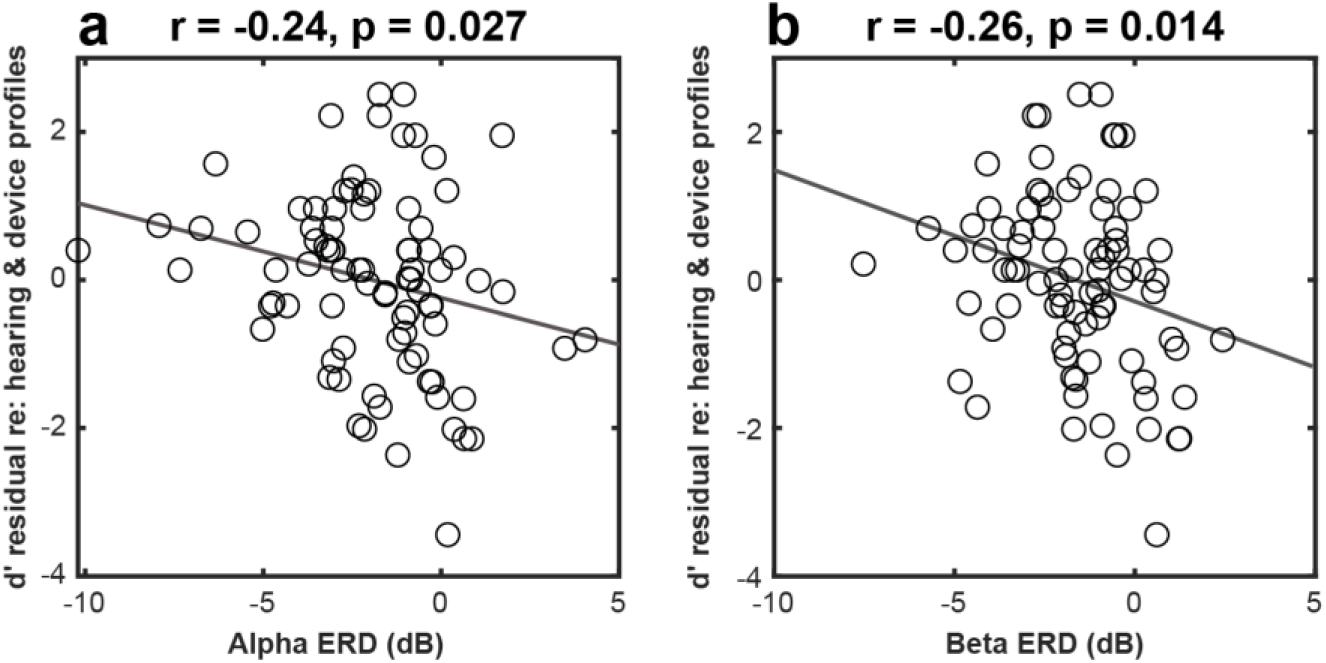
Relationship between event-related desynchronization (ERD) and the residual of d’ after regressing out hearing and device configurations (i.e., NHvSNHL, HAvCI, and AEvE contrasts). The correlation between d’ residual and alpha ERD is shown in panel **a**, and the correlation of d’ residual with beta ERD is illustrated in panel **b**.

In our sample of listeners with SNHL, the onset of hearing loss and the age at which hearing loss intervention was received differed across the groups (Table 1). As a post-hoc analysis, we ran a separate multiple regression model, with the HAvCI contrast, AEvE contrast, hearing loss onset, and age of receiving intervention as predictor variables, to explore whether these variables predicted the accuracy of the task. A full description of this model is provided in Supplementary 3. Briefly, the model showed that neither the onset of hearing loss nor the age at hearing loss intervention predicted performance on the task (*p* > 0.05), indicating that neither factor affects the detectability of auditory objects in listeners with SNHL.

## 4. Discussion

This study sought to investigate the cortical correlates of auditory object detection in subjects with different forms of hearing ability and listening configurations. There were several main findings. First, all groups performed the task well above chance. However, CI users, regardless of whether they had functional acoustic hearing, performed significantly worse than the NH and A-only groups. Second, evoked responses to the emergence of the auditory object were weaker in listeners with HA than NH listeners and absent in CI users (both A+E and E-only groups). Third, while delta and theta ERS following the auditory object were present in the NH group, neither of these responses was present in listeners with SNHL, except for the A+E group, which showed a delta ERS to the auditory object. Fourth, contrary to low-frequency ERSP, alpha and beta ERD responses elicited by the emergence of auditory objects were comparable across groups. Finally, regression modelling revealed that SFG task accuracy was predicted by both hearing configurations and cortical neural activity (indexed by alpha and beta ERD).

### 4.1 The ability to detect auditory objects is degraded in CI users

As hypothesized, listeners with SNHL performed worse in the SFG task relative to NH individuals. Previous studies have shown that SNHL impairs SiN perception (Dubno et al., 1984; Smith et al., 2024; Smoorenburg, 1992) and auditory stream segregation (Boncz et al., 2024; Grimault et al., 2001; Hong & Turner, 2006). In line with these findings, our study results indicate that the ability to detect auditory objects embedded in background noise is diminished by SNHL. Consistent with our hypothesis, this ability is even more degraded in CI users compared to listeners with SNHL who do not use a CI (i.e., A-only listeners), as planned contrast analysis showed significant differences in d’ between the CI and HA users (HAvCI contrast; Table 3). These results could be explained by the fact that A-only subjects retained some residual spectrotemporal processing, while CI users relied on their devices, which limits spectrotemporal processing of sounds (Anderson et al., 2011; Bierer et al., 2015; Goehring et al., 2019), affecting the detection of auditory objects in CI users.

Contrary to our hypothesis, we did not find significant differences between CI users with and without functional acoustic hearing for task performance (Table 3, AEvE contrast). This suggests that the presence of functional acoustic hearing in the unimplanted ear does not seem to contribute to the detectability of auditory objects in the SFG paradigm. These findings contradict previous reports showing a superior performance on SiN tasks in bilateral CI users relative to bimodal CI users (Blamey et al., 2015; Schafer et al., 2011). In our study, A+E participants had severe to profound hearing thresholds in the frequency range of 1000 to 8000 Hz for both the unaided non-CI and CI ears (Figure 2c). These hearing levels were similar to the unaided CI thresholds of the E-only group (Figure 2d). Specifically, the thresholds were above the presentation level of the stimulus (i.e., 70 dB SPL), meaning that the unaided hearing in both groups was functionally similar for the experimental stimulus. Given that the stimuli used for the SFG paradigm were limited to the electric hearing frequency range (1000 to 8000 Hz), having poor unaided hearing thresholds in both CI groups might have resulted in similar performance in the SFG task between the two groups. However, the A+E group had better low-frequency (< 1000 Hz) acoustic hearing than the E-only group, which might influence auditory grouping using a stimulus design with auditory objects composed of low-frequency tones. Future studies examining the effect of different hearing ranges on auditory object detection are needed.

The results of this study showed that response bias was similar across the groups. The measured criterion (c) had a negative value for all groups, indicating a liberal response bias, i.e., listeners were more likely to report that the auditory object was detected, even when a “Ground” trial was presented. These results suggest that the presence of SNHL does not influence the response strategy in the SFG task.

With respect to reaction time, no differences were observed concerning RT between groups. It is important to remind the reader that we designed the task so that the participants had to respond after the stimulus offset. Thus, this task design does not allow us to properly investigate how much information must accumulate before an object is detected; rather, any differences in RT are more likely to reflect uncertainty in the decision stage. Thus, our findings indicate that listeners with SNHL may take more time to decide whether they detected the emergence of auditory objects.

### 4.2 Auditory object-related evoked responses

In the current study, a P1-N1-P2-N2 complex was elicited following the figure onset, but this neural signature was only observed in the NH group. The topography of this complex showed a frontocentral distribution, which resembles the stimulus onset response. However, the topography also resembles the ORN response, which has been proposed to reflect processes involved in auditory figure-ground segregation (Winkler & Denham, 2024). In this study, we did not observe the ORN, P400, or the slow drift in the evoked response, which have been reported in previous studies that used the SFG paradigm (Boncz et al., 2024; Guo et al., 2022; Teki et al., 2016; Tóth et al., 2016a). The absence of these responses can be attributed to the usage of a 2-Hz high-pass filter to remove CI-related artifacts, which eliminates the possibility of seeing these responses. Although we did not find ORN responses in the current study, we cannot rule out that the neural generators of the ORN contributed to the responses here.

Listeners with SNHL showed attenuated evoked responses to the auditory object emergence relative to NH individuals, which is in line with our hypothesis. In particular, the A-only group showed visibly weaker evoked responses following the onset of the transition compared to the NH group; however, these evoked responses were characterized by a visible N1 component, which differs from the evoked response complex seen in the NH group (Figure 4a). These findings are compatible with previous reports showing degraded phase-locking responses to speech presented in a background of noise in listeners with SNHL (Ananthakrishnan et al., 2016; Henry & Heinz, 2013; Mai & Howell, 2023; Nash-Kille & Sharma, 2014; Woolf et al., 1981).

However, the current study results showed a lack of neural phase-locking responses to auditory object emergence in CI users. This conclusion was based on (1) the absence of visible evoked responses to the auditory object onset in the A+E and E-only groups (Figure 4a) and (2) the lack of significant ITPC clusters for both of these groups. To our knowledge, this is the first study that explored the neural correlates of the SFG paradigm in the CI population. Numerous studies have shown that CI users exhibit evoked responses to speech presented in a background of noise (Berger et al., 2023; Burkhardt et al., 2022; Finke et al., 2016; Shim et al., 2023; Soshi et al., 2014). Accordingly, the lack of evoked responses to auditory object emergence for CI users in the current study is surprising. One explanation for this observation is that the degree of SNHL severity might have suppressed neural phase-locking responses. Nash-Kille and Sharma (2014) found that the ITPC level decreases as SNHL increases in a sample of children with different degrees of SNHL, despite using aided hearing to listen to speech stimuli. Therefore, it could be possible that the absence of evoked responses to auditory object emergence in this study is due to the profound hearing loss levels in our CI sample. Alternatively, the lack of auditory-object-related evoked responses in the CI groups could be explained by the spectral and temporal processing limitations imposed by the CI device. Factors, such as physical placement of the CI in the cochlea (Chatterjee et al., 2006; Cooper & Roberts, 2007) and number of CI channels (Fu & Nogaki, 2005; Henry & Turner, 2003), can limit spectral resolution. In addition, CI listeners often exhibit degraded temporal resolution abilities, reflected as worse performance on gap detection (Cesur & Derinsu, 2020; Tuz et al., 2020) and amplitude modulation discrimination tasks (Shannon, 1992; Won et al., 2015) relative to NH individuals. Detecting auditory objects in the SFG paradigm fundamentally relies on the ability to detect rapid changes in the spectral features of the stimulus over time (Teki et al., 2013). These spectral and temporal limitations of the CI would lead to inconsistent timing of phase-locked neural responses impacting the detectability of auditory objects and making this process more variable.

Surprisingly, listeners with SNHL had a high accuracy level on the task despite having weaker or non-existent evoked responses to auditory object emergence. This indicates that evoked responses elicited following the transition onset were not a required condition for auditory object formation. Rather, induced spectral power indices of auditory object detection served as a better predictor applied across a broader range of hearing configurations. The temporal-coherence-based auditory streaming theory (Elhilali et al., 2009; Lu et al., 2017; Shamma et al., 2011; Teki et al., 2011), which asserts that auditory objects build up over time when their spectral features remain unchanged, may explain these results. It is possible that listeners with SNHL relied on non-time-locked, induced cortical activity to perform the task.

### 4.3 Delta and theta ERS magnitudes during auditory object emergence are attenuated in listeners with SNHL

Our study shows a lack of delta and theta ERS to the auditory object in all listeners with SNHL, except for the A+E group, which showed a delta ERS. In auditory scene analysis, low-frequency (delta and theta) power has been linked to neural entrainment to target sounds embedded within a noisy background (Ding & Simon, 2012; Ng et al., 2012; Riecke et al., 2015) and multiple-stream segregation (Tóth et al., 2016b, 2020). Furthermore, delta and theta ERS have been found to increase when attending to auditory streams (Ding & Simon, 2012; Golumbic et al., 2013; Lakatos et al., 2013; Tóth et al., 2016b). In the current study, the observed delta and theta ERS might be associated with auditory-object-related phase-locking responses, or with attending to auditory objects, as the current study required active participation in the task.

The permutation analysis showed significant clusters for both delta and theta ERS only in the NH group (Table 2). The time range of such clusters overlapped with the latency of object onset-related evoked responses. In addition, topographical mapping of delta and theta ERS activity in the NH group showed a scalp distribution similar to that of the ITPC (Figure 6). The ITPC topography resembles the typical evoked N1 topography (Alcaini et al., 1994), showing a frontocentral focus. As evoked responses were not subtracted from the ERSP analysis, it is likely that delta and theta ERS activity, seen in the time-frequency results following the transition onset, reflects a contribution from auditory cortical evoked responses that commonly show up in the frontocentral electrodes (Alcaini et al., 1994).

### 4.4 The magnitude of alpha and beta ERD activity during auditory object emergence is comparable across groups

We found that neither alpha nor beta desynchronization elicited by the presentation of the auditory object differed across groups. These results indicate that high-frequency oscillations are not clearly affected by SNHL, as is the case for low-frequency induced spectral power, and that oscillations in the higher frequency range may reflect cortical cognitive signals. Functionally, alpha modulation has been proposed to play a role in inhibition, where alpha ERD is thought to reflect a state of suppression of task-irrelevant information (Jensen & Mazaheri, 2010). In line with this view, the alpha ERD seen following the transition onset may be linked to the process of suppressing the background noise to enhance the detection of auditory objects. In addition, modulations in alpha oscillations have been postulated as a mechanism of attentional suppression (for review, see Foxe & Snyder, 2011, and Weisz et al., 2011). In a study where digits were presented in noise, beta and alpha ERD appeared when subjects actively listened to the auditory signals but not when they were passively listening to the stimuli (Dimitrijevic et al., 2017). As all subjects in the current study were required to actively attend and look for auditory objects, the suppression of alpha and beta oscillations may reflect central attentional modulation. Furthermore, beta oscillations have been linked to predictive mechanisms (Arnal, 2012; Arnal & Giraud, 2012; Bastiaansen et al., 1999; Sedley et al., 2016; Todorovic et al., 2015). In particular, beta oscillations have been shown to occur as a result of prediction changes, i.e., beta activity was involved in updating the prediction content (Sedley et al., 2016). According to these views, the emergence of beta ERD following the transition onset may be a marker for updating the predictions from random to coherent presentation of tone pips. In the context of pitch deviance, Chang et al. (2018) found larger beta ERD responses to predictable pitch tones compared to unpredictable (deviant) pitch tones. Thus, it might be possible that beta desynchronization is induced as an effect of the predictable upcoming changes in the stimulus spectral features, which occur during the transition period. Overall, the data from our study suggests that alpha and beta oscillations are likely related to neural cognitive processing of auditory object detection.

Alpha ERD topographies show a wide distribution of activity covering the entire scalp, which was observed in all groups. However, alpha ERD was more densely localized in the frontal, central, parietal, and right temporal regions in the NH group. This alpha ERD topography is consistent with previous studies involving SiN tasks (Dimitrijevic et al., 2017; Obleser & Weisz, 2012; Paul et al., 2021). For example, Dimitrijevic et al. (2017) reported that when participants were actively listening to SiN stimuli, alpha ERD was present in central and parietal regions. Additionally, a frontocentral topography of alpha ERD was found by Paul et al. (2021), wherein alpha ERD power was decreased as a function of effortful listening in the SiN task. Similarly, beta ERD topography showed widely diffuse activity covering the entire scalp, with more concentration in frontal and parietal regions seen in the NH group. The recruitment of parietal regions may be related to the involvement of brain areas responsible for auditory segregation, such as the intra-parietal sulcus (Teki et al., 2011, 2016). Overall, the findings from topographical mapping of alpha and beta ERD activity suggest the recruitment of widely distributed brain regions during the detection of auditory objects. However, future studies may benefit from using time-frequency source localization, or functional neuroimaging techniques to find the brain location of alpha/beta ERD activity.

### 4.5 SFG task accuracy is predicted by cortical activity beyond hearing configurations

In our regression modelling, the three contrasts were set as the main predictors, wherein NHvSNHL and HAvCI contrasts were significant (Table 4). When alpha and beta were added together to the regression model, the results showed they collectively accounted for significant new variance, even as neither were individually significant. However, when entered separately, alpha and beta were each significant predictors of task performance. These findings reflect the presence of a suppressor effect, meaning that the addition of both alpha and beta led to a reduction of their individual predictive power. These results cumulatively suggest that alpha and beta oscillations have a similar role in auditory object detection. It is important to mention that we did not design the current study to dissociate the contribution of alpha and beta to the detectability of auditory objects. As mentioned previously (section 4.4), alpha oscillations might be involved in recruiting attentional resources (Dimitrijevic et al., 2017) or suppressing task-irrelevant stimuli (Jensen & Mazaheri, 2010), whereas beta oscillations might be linked to predictive processes (Arnal, 2012; Arnal & Giraud, 2012; Bastiaansen et al., 1999; Sedley et al., 2016; Todorovic et al., 2015). The fact that these were so highly correlated suggests these processes might be linked—active suppression of the ground (reflected in alpha ERD) enables better prediction of the continuation of the figure (reflected in beta ERD). Future investigations looking into the differential role of alpha and beta in auditory object detection are needed.

Interestingly, in all of the performed regression models, only NHvSNHL and HAvCI contrasts significantly predicted the performance on the SFG task. These results indicate that SFG task performance is affected by the presence of SNHL, and among listeners with SNHL, object detection is influenced by whether an individual uses a CI or HA. Notably, we did not find a significant difference between CI users with and without functional acoustic hearing, which suggests that low-frequency acoustic hearing does not contribute to auditory object detectability in the CI population. This might be because the unaided hearing levels were poor in our A+E participants for the non-CI ear. Whether less severe functional acoustic hearing on the unimplanted ear improves auditory object detection remains an open question for future research.

In the current study, listeners with SNHL had a high variance concerning SNHL onset and the age of receiving intervention for SNHL (Table 1). Our analysis showed that neither of these factors predicted the SFG task accuracy (Supplementary 3). Notably, these results are consistent with previous studies showing that, for post-lingual adults with SNHL, the age of receiving cochlear implantation and the duration of deafness did not impact SiN outcomes (Herzog et al., 2003; Olze et al., 2012; Távora-Vieira et al., 2015). Given that our sample included adults with post-lingual SNHL, differences in the SNHL onset and age at intervention do not have a strong influence on auditory object detectability.

### 4.6 Limitations and Future Directions

The current study has several limitations that should be noted. Given that we removed many ICA components in the CI groups (as they likely reflected an artifact of the electrical stimulation of the CI), this could have influenced the results in this study. However, we believe that the absence of auditory-object related phase-locking responses in the A+E and E-only groups is not a result of CI-related artifact removal for several reasons. First, despite the low number of ICA components removed in the A-only group, phase-locking to the transition onset was very weak, as evident in the evoked response and ITPC results (Figure 4). Second, both the E-only and A+E groups exhibited evoked and ITPC responses at the onset and offset of the sound. However, it remains possible that evoked responses were attenuated in the E-only and A+E groups due to CI-related artifact removal.

The coherence level, defined as the number of tonal elements that become spectrally coherent over time following the transition onset, of our SFG stimuli was fixed at six frequencies. For this study, we purposefully designed the Figure stimuli to be easily detectable for CI users, which was shown to result in high hit rates in previous studies done on NH listeners (Teki et al., 2011, 2013; Tóth et al., 2016a). Crucially, we did not manipulate the coherence level in this study, as the aim was to explore whether listeners with different auditory configurations can detect auditory objects. It is well-documented that the hit rate increases as a function of increasing coherence levels in the SFG paradigm (O’Sullivan et al., 2015; Teki et al., 2011, 2013). However, whether listeners with SNHL show improved auditory object detectability with increasing coherence levels in the SFG paradigm remains to be established.

The current study did not compare the neural response of the SFG paradigm between active and passive listening conditions. It has been previously shown that active listening to auditory objects in the SFG paradigm (as was done here) enhances behavioral performance and elicits stronger evoked responses than passive listening (O’Sullivan et al., 2015). Crucially, the study by Dimitrijevic et al. (2017) has shown that alpha and beta ERD emerged only when actively listening to SiN stimuli. Accordingly, alpha/beta ERD observed in this study may result from active listening to the SFG stimuli. Future studies comparing the magnitude of alpha/beta ERD activity between active and passive SFG listening conditions are warranted.

In this study, we did not compare the neural responses between hit and miss trials. Due to the high accuracy results observed in all groups, we could not investigate the neural correlates of hit versus miss trials. This was because we designed the SFG stimuli with a long duration (4 seconds) and high coherence level, making it easy for listeners with SNHL to perform the task. Notably, differences in neural responses between hit and miss trials of the SFG paradigm have been demonstrated by Tóth et al. (2016a), in which no evoked responses to the SFG stimulus were observed in the miss trials that were otherwise present on hit trials. Therefore, designing a more challenging task where more miss trials can be obtained from listeners with SNHL and looking into the neural correlates of hit and miss trials could be explored in future investigations.

Finally, given that CI users exhibit heterogeneous hearing profiles, multiple factors could influence auditory object detection. As noted previously, the spectrotemporal processing of the CI is dependent on the physical placement of and number of CI channels (Chatterjee et al., 2006; Cooper & Roberts, 2007; Fu & Nogaki, 2005; Henry & Turner, 2003). Moreover, the side of implantation may impact outcomes: users with right-sided CIs have demonstrated better performance on speech perception tasks compared to those with left-sided CIs (Henkin et al., 2014; Kraaijenga et al., 2018). Therefore, future studies should investigate how auditory object detection is influenced by CI-related factors such as electrode physical placement, number of CI channels, and side of implantation.

## 5. Conclusions

To our knowledge, this is the first study that directly compared neural correlates of auditory object detection among subjects with different auditory configurations: NH, A-only, A+E, and E-only. We found that listeners with SNHL had degraded cortical phase-locking of evoked responses to auditory objects despite performing well on the SFG task. Unique to CI users, no such evoked responses were observed. Delta and theta ERS responses during the auditory object emergence were diminished in SNHL groups, except for the A+E groups, which showed a delta ERS. However, auditory-object-related alpha and beta desynchronization was present in all groups to a similar extent. Hierarchical regression modelling revealed that alpha and beta desynchronization predicted the accuracy of the SFG task beyond peripheral hearing status. We, therefore, establish that optimal auditory object detection relies on both peripheral acuity and cortical processes.

## Data Availability

All data produced in the present study are available upon reasonable request to the authors.

## Acknowledgements

We would like to thank Laura Kiskunas for helping with data collection.

## CRediT authorship contribution statement

**Nour Alsabbagh**: Data curation, formal analysis, investigation, methodology, visualization, writing – original draft, writing – review and editing. **Bob McMurray**: Conceptualization, formal analysis, investigation, funding acquisition, project administration, methodology, formal analysis, writing – review and editing. **Timothy Griffiths**: Conceptualization, funding acquisition, project administration, methodology, writing – review and editing. **Joel Berger**: Methodology, writing – review and editing. **Kyogu Lee**: methodology, formal analysis, funding acquisition. **Phillip Gander**: Conceptualization, formal analysis, investigation, methodology, supervision, funding acquisition, project administration, writing – review and editing. **Inyong Choi**: Conceptualization, formal analysis, investigation, methodology, supervision, funding acquisition, project administration, writing – review and editing.

