## Supplementary materials for "Cortical oscillations predict auditory grouping in listeners with and without hearing loss"

### Supplementary 1

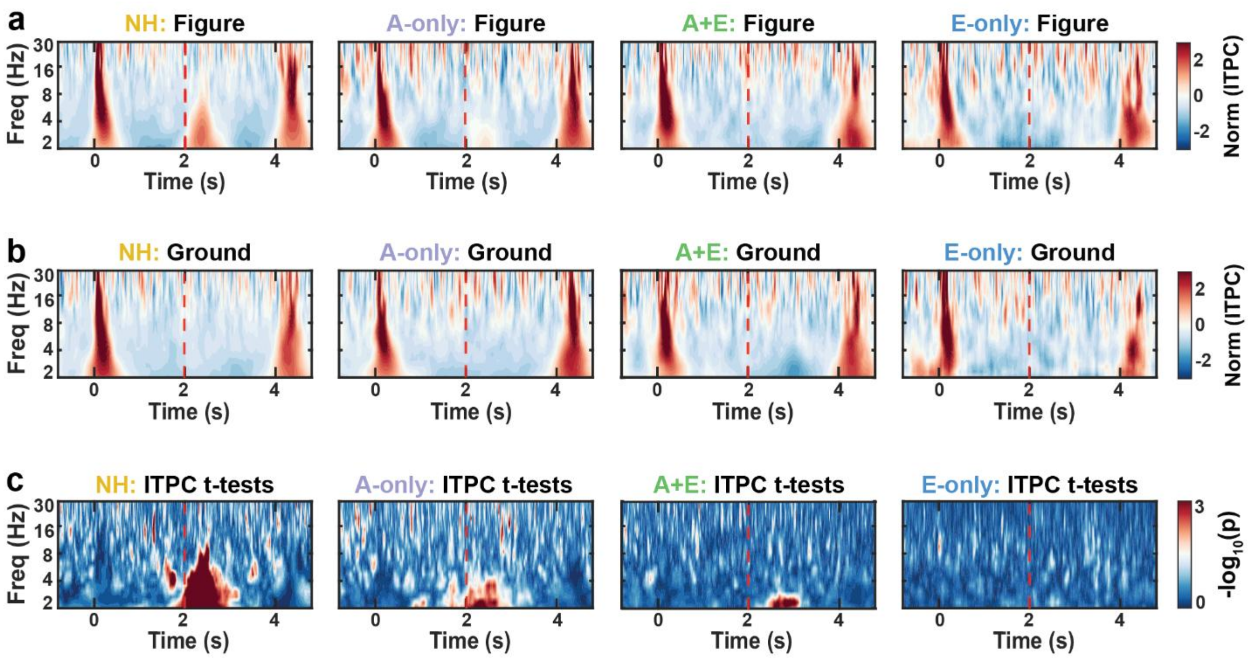


**Figure 1.** The intertrial phase coherence (ITPC) results. ITPC was measured by taking the average activity across frontocentral (Fz, FCz, Cz, FC1, and FC2) channels. Subsequently, ITPC results were normalized using z-score. Panel **a** shows group-level ITPC results for the Figure condition, while panel **b** shows ITPC results for the Ground. Positive ITPC values indicate a higher degree of phase coherence consistency across Figure trials, compared to the Ground condition. Conversely, negative ITPC values indicate a lower degree of phase coherence consistency across Figure trials. The statistical difference between each time-frequency ITPC bin is demonstrated in panel **c**, showing uncorrected paired-sample t-test results. The red scale color indicates a significant time-frequency point ($p < 0.05$). Vertical dashed red lines show the transition onset, i.e., the time of auditory object emergence.

### Supplementary 2

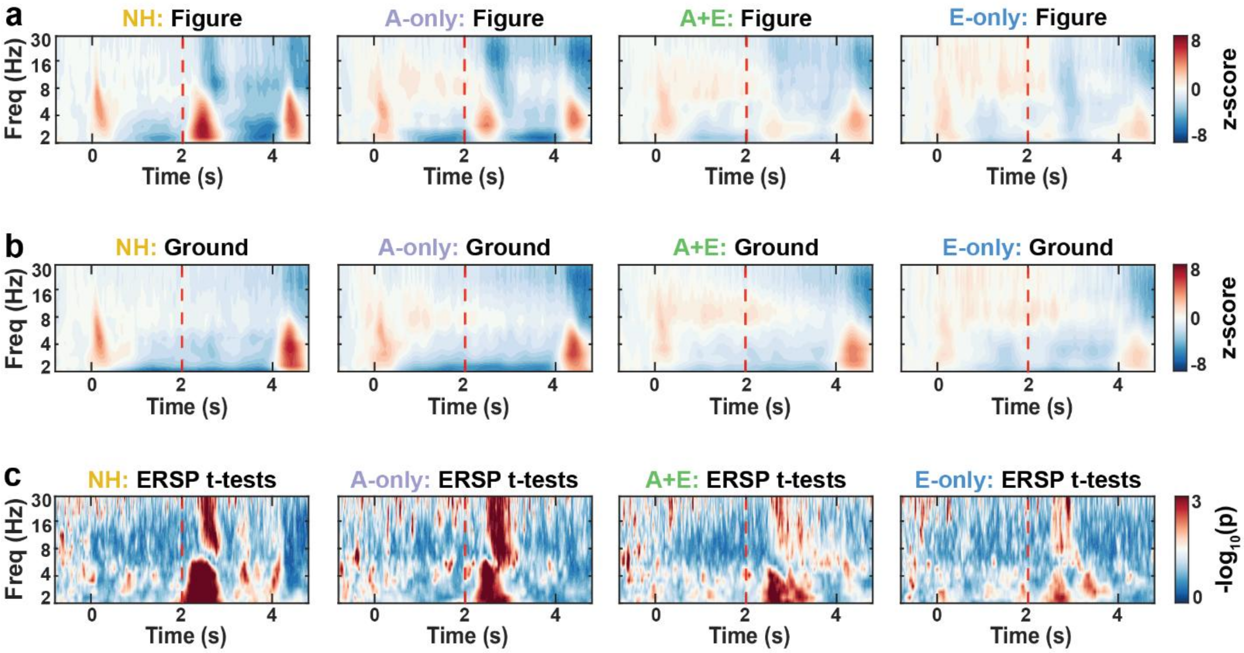


***Figure 2.*** *The event-related spectral perturbation (ERSP) results. ERSP was measured by averaging ERSP activity across the 50 remaining channels. The ERSP result for the Figure and Ground conditions is depicted in panels **a** and **b**, respectively. Panel **c** represents uncorrected paired sample t-tests comparing each time-frequency bin between the Figure and Ground conditions. Time-frequency points that are in red indicate significance (p < 0.05). Vertical dashed red lines show the transition onset, i.e., the time of auditory object emergence.*

**Supplementary 3**

In our sample of listeners with SNHL, the onset of hearing loss and the age at which participants received intervention for their hearing loss differed across the A-only, A+E, and E-only groups. In this supplementary analysis, we investigated whether these factors, i.e., SNHL onset (HL_onset) and age at hearing loss intervention (HL_interv), influence our results. We ran a multiple regression model with contrast 2 (HAvCI), contrast 3 (AEvE), HL_onset, and HL_interv as predictors (Contrast 1 was excluded from the analysis, given that both "HL" factors do not apply to the NH group), as follows:

$$d \sim HAvCI + AEvE + HL\_onset + HL\_interv \quad (1)$$

This model (1) was significant ($F_{4,58} = 5.023$, $p = 0.001$, $R^2 = 0.206$), wherein the HAvCI contrast was a significant predictor of d' ($p < 0.001$). A full summary of the multiple regression model is provided in Table 1. Importantly, neither HL_onset nor HL_interv predicted the detectability of auditory objects ($p > 0.05$) in listeners with SNHL. This analysis confirmed that hearing loss onset and the age at which the listener received an intervention did not have a significant effect on predicting performance on the task.

***Table 1.*** *Multiple regression model analysis results (N = 65)*

| **d'~ HAvCI + AEvE + HL_onset + HL_interv** | | | | |
|---|---|---|---|---|
| **predictors** | **β** | **SE** | **t value** | ***p* value** |
| Intercept | 0.037 | 0.117 | 0.322 | 0.788 |
| HAvCI | **0.582** | **0.207** | **2.813** | **0.006** |
| AEvE | 0.079 | 0.151 | 0.527 | 0.599 |
| HL_onset | -0.378 | 0.272 | -1.386 | 0.171 |
| HL_interv | 0.531 | 0.271 | 1.962 | 0.054 |
